# Age and gender profiles of HIV infection burden and viraemia: novel metrics for HIV epidemic control in African populations with high antiretroviral therapy coverage

**DOI:** 10.1101/2024.04.21.24306145

**Authors:** Andrea Brizzi, Joseph Kagaayi, Robert Ssekubugu, Lucie Abeler-Dörner, Alexandra Blenkinsop, David Bonsall, Larry W. Chang, Christophe Fraser, Ronald M. Galiwango, Godfrey Kigozi, Imogen Kyle, Mélodie Monod, Gertrude Nakigozi, Fred Nalugoda, Joseph G. Rosen, Oliver Laeyendecker, Thomas C. Quinn, M. Kate Grabowski, Steven J. Reynolds, Oliver Ratmann, the Rakai Health Sciences Program

## Abstract

**Introduction:** To prioritize and tailor interventions for ending AIDS by 2030 in Africa, it is important to characterize the population groups in which HIV viraemia is concentrating.

**Methods:** We analysed HIV testing and viral load data collected between 2013-2019 from the open, population-based Rakai Community Cohort Study (RCCS) in Uganda, to estimate HIV seroprevalence and population viral suppression over time by gender, one-year age bands and residence in inland and fishing communities. All estimates were standardized to the underlying source population using census data. We then assessed 95-95-95 targets in their ability to identify the populations in which viraemia concentrates.

**Results:** Following the implementation of Universal Test and Treat, the proportion of individuals with viraemia decreased from 4.9% (4.6%-5.3%) in 2013 to 1.9% (1.7%-2.2%) in 2019 in inland communities and from 19.1% (18.0%-20.4%) in 2013 to 4.7% (4.0%-5.5%) in 2019 in fishing communities. Viraemia did not concentrate in the age and gender groups furthest from achieving 95-95-95 targets. Instead, in both inland and fishing communities, women aged 25-29 and men aged 30-34 were the 5-year age groups that contributed most to population-level viraemia in 2019, despite these groups being close to or had already achieved 95-95-95 targets.

**Conclusions:** The 95-95-95 targets provide a useful benchmark for monitoring progress towards HIV epidemic control, but do not contextualize underlying population structures and so may direct interventions towards groups that represent a marginal fraction of the population with viraemia.

## 1 Introduction

The human immunodeficiency virus (HIV) continues to be a deadly infection. In 2022, there were an estimated 630,000 (480,000-880,000) deaths worldwide and 1.3 million new infections globally, of which more than half occurred in Eastern and Southern Africa (ESA) [1]. Combination antiretroviral treatment (ART) effectively suppresses HIV viraemia and is highly effective in reducing HIV-associated mortality[2–4] and preventing onward HIV transmission[5,6]. Consequently, treatment has been adopted as a public health strategy to prevent new HIV infections (treatment as prevention, TasP)[1,7,8]. Progress in the implementation of TasP programs is primarily measured through the Joint United Nations Programme on HIV/AIDS (UNAIDS) “95-95-95” targets, with the aim that 95% of people living with HIV (PLHIV) are aware of their status, 95% of those aware are on antiretroviral therapy, and 95% of those on ART achieve viral suppression. All countries in ESA have committed to achieving these targets by 2025[9], as part of the UN Sustainable Development Goals[10,11].

With the expansion of HIV testing and treatment services, many countries in ESA have either attained or are close to attaining the “95-95-95” targets at a population level[12,13]. Thus, determining novel effective methods to measure HIV epidemic control and direct future efforts to accelerate epidemic decline in such contexts is required. While stratification of “95-95-95” targets by age and gender represents one approach, current evidence suggests that these metrics may not be predictive of incidence declines[14–17]. Although populations stratifications of existing targets are useful in highlighting important inequalities in treatment coverage[18], they do not account for underlying demographic structures or distributions of HIV burden in the population. Consequently, they may not provide a complete understanding of the people most at risk of HIV-related morality and onward transmission, which is necessary to tailor and implement effective prevention interventions.

In this study, we consider an alternative metric for assessing HIV control, one that integrates demographic information, specifically the age and gender profiles of the population burdens of HIV infection and HIV viraemia. Using longitudinal data from the Rakai Community Cohort Study (RCCS), a large population-based HIV surveillance cohort in southern Uganda, we compare the proposed profiles against the HIV seroprevalence and the prevalence of viraemia among PLHIV implied by the “95-95-95” targets. Our analyses were conducted across two distinct geographic settings with unique demographic compositions, including agrarian villages and semi-urban trading centres with population pyramids typical of most ESA communities and hyper endemic Lake Victoria fishing communities with atypical population structures. The study timeframe, spanning 2013 to 2019, coincided with the national expansion of ART programs.

## 2 Methods

### 2.1 Study population

The RCCS is a population-based cohort of individuals aged 15 to 49 years established in 1994 by the Rakai Health Sciences Program (RHSP). The RCCS is implemented in communities in and around Rakai District[19], bordering Lake Victoria to the east and Tanzania to the south. Between July 2013 and May 2019, four survey rounds (survey rounds 16, 17, 18 and 19) were conducted in four Lake Victoria fishing and 36 inland agrarian and semi-urban communities. RCCS survey methods have been described previously[20]. In brief, at each survey round, the RCCS conducted a household census and subsequently invited all individuals aged 15-49 years and residing in the communities with intention to stay for at least 1 month to participate. Consent was obtained privately by a trained RCCS interviewer. Following consent, participants were interviewed in a private location, typically a tent, for demographic characteristics as well as behavioral and healthcare behavior and asked to provide a blood sample for HIV testing and future laboratory studies. All participants were offered pre-test and post-test counseling, and referrals to ART services if living with HIV.

### 2.2 Population sizes

Age and gender-specific population sizes in each survey round were estimated from line-list data of census-eligible individuals that were reported by household heads during the census. The reported ages tended to reflect grouping patterns around multiples of 5, which we disaggregated by smoothing population sizes over age (Supplementary Text S1.1 and Supplementary Figure S2[21]).

### 2.3 Measurement of HIV serostatus and viraemia

All RCCS participants were offered free HIV testing. Testing was performed through a combination of three rapid tests with confirmation of positive, faintly positive, and discordant results by at least two EIAs and Western Blot or DNA PCR[22]. Overall, 99.7% of participants accepted HIV testing across survey rounds. Among participants living with HIV, HIV-1 viral load was measured on stored serum samples using the Abbott real-time m2000 assay (Abbott Laboratories, IL, USA). Overall, viral load measurements were obtained from nearly all (99.4%) participants living with HIV [23–25]. Viral suppression was defined as a viral load measurement below 1,000 HIV RNA copies/mL plasma, following recommendations of the World Health Organization (WHO)[26] at the time. People who did not achieve viral suppression were defined as viraemic. From these data, we first estimated the population-level HIV seroprevalence (i.e., probability for an individual to be living with HIV) with a non-parametric Bayesian model over the age of individuals stratifying by gender, survey round and community type (i.e., fishing vs. inland), using the data from first-time participants as proxy for non-participating individuals (Supplementary Text, S1.2)[21]. Then, the same model was used to estimate the proportion of individuals exhibiting viraemia among people living with HIV (Supplementary TextS1.3) and among all individuals (Supplementary Text S1.4).

### 2.4. HIV infection burden and burden of viraemia profiles

We further characterized the HIV burden profile, i.e., the contribution of each gender and age subgroup to the total number of PLHIV. First, we estimated the number *I*_*g,a*_ of individuals of gender *g* and age *a* living with HIV in the population by multiplying the estimated age and gender specific seroprevalence of HIV 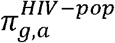 with the estimated number of census-eligible individuals *N*_*g,a*_, specifically 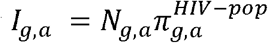. We then computed the HIV burden profile by taking the ratio of the number *I*_*g,a*_ of individuals of gender *g* and age *a* living with HIV against the sum over all age and gender groups:

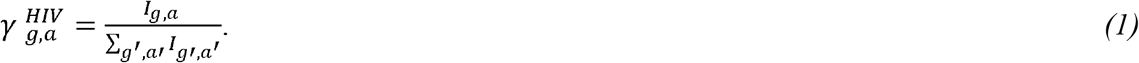

We analogously described the burden profile of HIV viraemia, i.e., the contribution of each gender and age subgroup to the total number of individuals exhibiting viraemia. First, we estimated the number *V*_*g,a*_ of individuals of gender *g* and age *a* exhibiting viraemia by multiplying the age and gender specific prevalence of viraemia 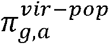 by *N*_*g,a*_ the estimated number of census-eligible individuals *N*_*g,a*_, specifically 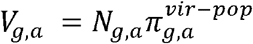. We then computed the profile of viraemia by taking the ratio of the number *V*_*g,a*_ of individuals of gender *g* and age *a* against the sum over all age and gender groups:

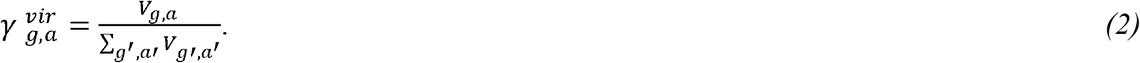

### 2.5 Ethics

The study was independently reviewed and approved by the Ugandan Virus Research Institute, Research and Ethics Committee, protocol GC/127/13/01/16; the Ugandan National Council for Science and Technology; and the Western Institutional Review Board, protocol 200313317. All study participants provided written informed consent at baseline and follow-up visits using institutional review board approved forms. This project was reviewed in accordance with CDC human research protection procedures and was determined to be research, but CDC investigators did not interact with human subjects or have access to identifiable data or specimens for research purposes. RCCS participants received 10,000UGX (approximately 2.50USD) in compensation for their time and costs to participate in each survey.

## 3 Results

### 3.1 Age and gender burden profiles of HIV infection

From July 2013 to May 2019, across the four survey rounds, a total of 37,743 individuals participated in the RCCS at least once (see Supplementary Table S1 for participation rates and Supplementary Figure S1 for study flowchart). Of these, 22,215 (58.86%) participated for the first time (Supplementary Table S2). Inland communities were characterized by expansive population pyramids with a predominantly young population, consistent with rapid population growth across Uganda and ESA[27]. In contrast, fishing communities were characterized by constrictive population pyramids comprising primarily working-age adults (20-40 years) and substantially fewer children or families than in inland communities (Figure 1b). Gender distributions also differed, with women accounting for 52.2% of the population in inland communities and 44.3% in fishing communities (Table 1).

**Table 1.**
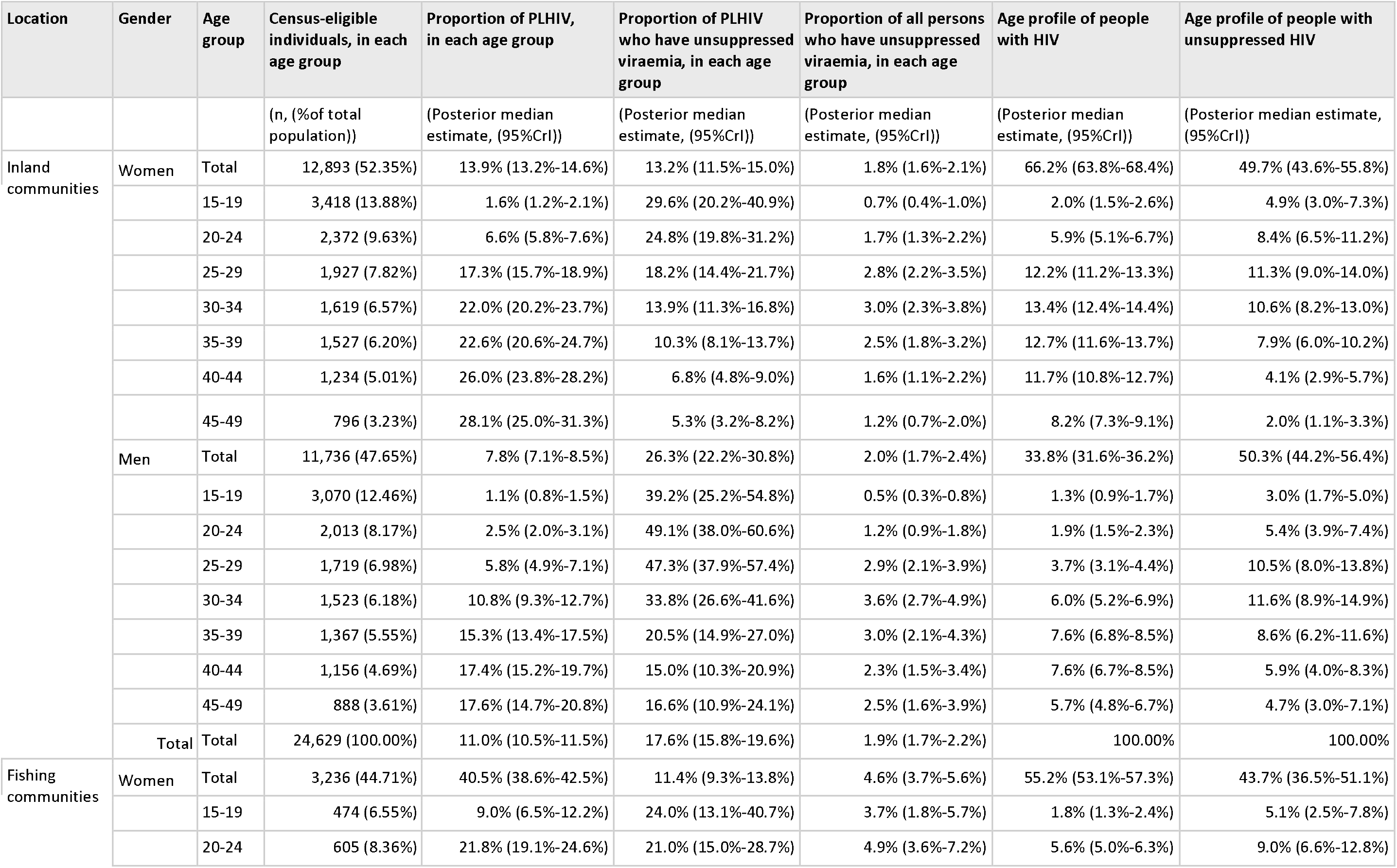
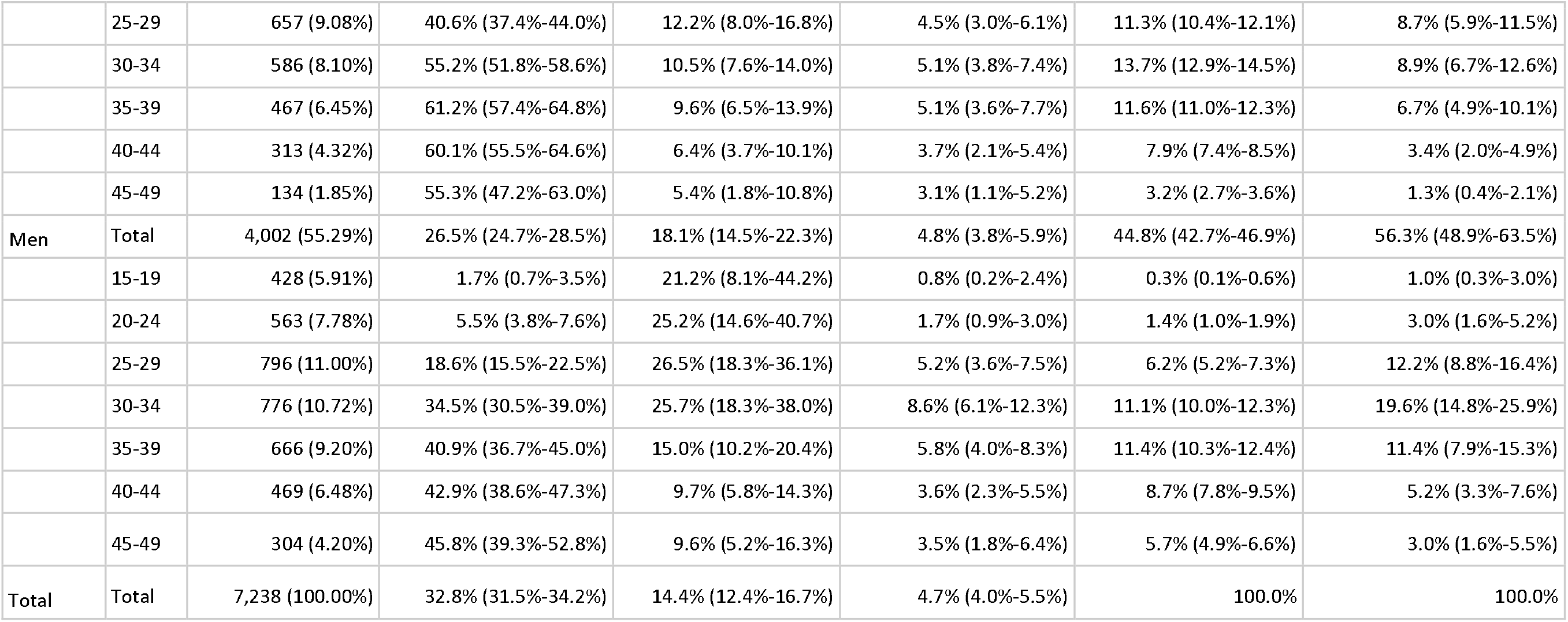
Characteristics of populations with and without HIV in the Rakai Community Cohort Study, June 2018-May 2019 (round 19)

| Location | Gender | Age group | Census-eligible individuals, in each age group | Proportion of PLHIV, in each age group | Proportion of PLHIV who have unsuppressed viraemia, in each age group | Proportion of all persons who have unsuppressed viraemia, in each age group | Age profile of people with HIV | Age profile of people with unsuppressed HIV |
| --- | --- | --- | --- | --- | --- | --- | --- | --- |
|  |  |  | (n, (% of total population)) | (Posterior median estimate, (95%CrI)) | (Posterior median estimate, (95%CrI)) | (Posterior median estimate, (95%CrI)) | (Posterior median estimate, (95%CrI)) | (Posterior median estimate, (95%CrI)) |
| Inland communities | Women | Total | 12,893 (52.35%) | 13.9% (13.2%-14.6%) | 13.2% (11.5%-15.0%) | 1.8% (1.6%-2.1%) | 66.2% (63.8%-68.4%) | 49.7% (43.6%-55.8%) |
|  |  | 15-19 | 3,418 (13.88%) | 1.6% (1.2%-2.1%) | 29.6% (20.2%-40.9%) | 0.7% (0.4%-1.0%) | 2.0% (1.5%-2.6%) | 4.9% (3.0%-7.3%) |
|  |  | 20-24 | 2,372 (9.63%) | 6.6% (5.8%-7.6%) | 24.8% (19.8%-31.2%) | 1.7% (1.3%-2.2%) | 5.9% (5.1%-6.7%) | 8.4% (6.5%-11.2%) |
|  |  | 25-29 | 1,927 (7.82%) | 17.3% (15.7%-18.9%) | 18.2% (14.4%-21.7%) | 2.8% (2.2%-3.5%) | 12.2% (11.2%-13.3%) | 11.3% (9.0%-14.0%) |
|  |  | 30-34 | 1,619 (6.57%) | 22.0% (20.2%-23.7%) | 13.9% (11.3%-16.8%) | 3.0% (2.3%-3.8%) | 13.4% (12.4%-14.4%) | 10.6% (8.2%-13.0%) |
|  |  | 35-39 | 1,527 (6.20%) | 22.6% (20.6%-24.7%) | 10.3% (8.1%-13.7%) | 2.5% (1.8%-3.2%) | 12.7% (11.6%-13.7%) | 7.9% (6.0%-10.2%) |
|  |  | 40-44 | 1,234 (5.01%) | 26.0% (23.8%-28.2%) | 6.8% (4.8%-9.0%) | 1.6% (1.1%-2.2%) | 11.7% (10.8%-12.7%) | 4.1% (2.9%-5.7%) |
|  |  | 45-49 | 796 (3.23%) | 28.1% (25.0%-31.3%) | 5.3% (3.2%-8.2%) | 1.2% (0.7%-2.0%) | 8.2% (7.3%-9.1%) | 2.0% (1.1%-3.3%) |
|  | Men | Total | 11,736 (47.65%) | 7.8% (7.1%-8.5%) | 26.3% (22.2%-30.8%) | 2.0% (1.7%-2.4%) | 33.8% (31.6%-36.2%) | 50.3% (44.2%-56.4%) |
|  |  | 15-19 | 3,070 (12.46%) | 1.1% (0.8%-1.5%) | 39.2% (25.2%-54.8%) | 0.5% (0.3%-0.8%) | 1.3% (0.9%-1.7%) | 3.0% (1.7%-5.0%) |
|  |  | 20-24 | 2,013 (8.17%) | 2.5% (2.0%-3.1%) | 49.1% (38.0%-60.6%) | 1.2% (0.9%-1.8%) | 1.9% (1.5%-2.3%) | 5.4% (3.9%-7.4%) |
|  |  | 25-29 | 1,719 (6.98%) | 5.8% (4.9%-7.1%) | 47.3% (37.9%-57.4%) | 2.9% (2.1%-3.9%) | 3.7% (3.1%-4.4%) | 10.5% (8.0%-13.8%) |
|  |  | 30-34 | 1,523 (6.18%) | 10.8% (9.3%-12.7%) | 33.8% (26.6%-41.6%) | 3.6% (2.7%-4.9%) | 6.0% (5.2%-6.9%) | 11.6% (8.9%-14.9%) |
|  |  | 35-39 | 1,367 (5.55%) | 15.3% (13.4%-17.5%) | 20.5% (14.9%-27.0%) | 3.0% (2.1%-4.3%) | 7.6% (6.8%-8.5%) | 8.6% (6.2%-11.6%) |
|  |  | 40-44 | 1,156 (4.69%) | 17.4% (15.2%-19.7%) | 15.0% (10.3%-20.9%) | 2.3% (1.5%-3.4%) | 7.6% (6.7%-8.5%) | 5.9% (4.0%-8.3%) |
|  |  | 45-49 | 888 (3.61%) | 17.6% (14.7%-20.8%) | 16.6% (10.9%-24.1%) | 2.5% (1.6%-3.9%) | 5.7% (4.8%-6.7%) | 4.7% (3.0%-7.1%) |
|  | Total | Total | 24,629 (100.00%) | 11.0% (10.5%-11.5%) | 17.6% (15.8%-19.6%) | 1.9% (1.7%-2.2%) | 100.00% | 100.00% |
| Fishing communities | Women | Total | 3,236 (44.71%) | 40.5% (38.6%-42.5%) | 11.4% (9.3%-13.8%) | 4.6% (3.7%-5.6%) | 55.2% (53.1%-57.3%) | 43.7% (36.5%-51.1%) |
|  |  | 15-19 | 474 (6.55%) | 9.0% (6.5%-12.2%) | 24.0% (13.1%-40.7%) | 3.7% (1.8%-5.7%) | 1.8% (1.3%-2.4%) | 5.1% (2.5%-7.8%) |
|  |  | 20-24 | 605 (8.36%) | 21.8% (19.1%-24.6%) | 21.0% (15.0%-28.7%) | 4.9% (3.6%-7.2%) | 5.6% (5.0%-6.3%) | 9.0% (6.6%-12.8%) |

|  |  |  |  |  |  |  |  |
| --- | --- | --- | --- | --- | --- | --- | --- |
|  | 25-29 | 657 (9.08%) | 40.6% (37.4%-44.0%) | 12.2% (8.0%-16.8%) | 4.5% (3.0%-6.1%) | 11.3% (10.4%-12.1%) | 8.7% (5.9%-11.5%) |
|  | 30-34 | 586 (8.10%) | 55.2% (51.8%-58.6%) | 10.5% (7.6%-14.0%) | 5.1% (3.8%-7.4%) | 13.7% (12.9%-14.5%) | 8.9% (6.7%-12.6%) |
|  | 35-39 | 467 (6.45%) | 61.2% (57.4%-64.8%) | 9.6% (6.5%-13.9%) | 5.1% (3.6%-7.7%) | 11.6% (11.0%-12.3%) | 6.7% (4.9%-10.1%) |
|  | 40-44 | 313 (4.32%) | 60.1% (55.5%-64.6%) | 6.4% (3.7%-10.1%) | 3.7% (2.1%-5.4%) | 7.9% (7.4%-8.5%) | 3.4% (2.0%-4.9%) |
|  | 45-49 | 134 (1.85%) | 55.3% (47.2%-63.0%) | 5.4% (1.8%-10.8%) | 3.1% (1.1%-5.2%) | 3.2% (2.7%-3.6%) | 1.3% (0.4%-2.1%) |
| Men | Total | 4,002 (55.29%) | 26.5% (24.7%-28.5%) | 18.1% (14.5%-22.3%) | 4.8% (3.8%-5.9%) | 44.8% (42.7%-46.9%) | 56.3% (48.9%-63.5%) |
|  | 15-19 | 428 (5.91%) | 1.7% (0.7%-3.5%) | 21.2% (8.1%-44.2%) | 0.8% (0.2%-2.4%) | 0.3% (0.1%-0.6%) | 1.0% (0.3%-3.0%) |
|  | 20-24 | 563 (7.78%) | 5.5% (3.8%-7.6%) | 25.2% (14.6%-40.7%) | 1.7% (0.9%-3.0%) | 1.4% (1.0%-1.9%) | 3.0% (1.6%-5.2%) |
|  | 25-29 | 796 (11.00%) | 18.6% (15.5%-22.5%) | 26.5% (18.3%-36.1%) | 5.2% (3.6%-7.5%) | 6.2% (5.2%-7.3%) | 12.2% (8.8%-16.4%) |
|  | 30-34 | 776 (10.72%) | 34.5% (30.5%-39.0%) | 25.7% (18.3%-38.0%) | 8.6% (6.1%-12.3%) | 11.1% (10.0%-12.3%) | 19.6% (14.8%-25.9%) |
|  | 35-39 | 666 (9.20%) | 40.9% (36.7%-45.0%) | 15.0% (10.2%-20.4%) | 5.8% (4.0%-8.3%) | 11.4% (10.3%-12.4%) | 11.4% (7.9%-15.3%) |
|  | 40-44 | 469 (6.48%) | 42.9% (38.6%-47.3%) | 9.7% (5.8%-14.3%) | 3.6% (2.3%-5.5%) | 8.7% (7.8%-9.5%) | 5.2% (3.3%-7.6%) |
|  | 45-49 | 304 (4.20%) | 45.8% (39.3%-52.8%) | 9.6% (5.2%-16.3%) | 3.5% (1.8%-6.4%) | 5.7% (4.9%-6.6%) | 3.0% (1.6%-5.5%) |
| Total | Total | 7,238 (100.00%) | 32.8% (31.5%-34.2%) | 14.4% (12.4%-16.7%) | 4.7% (4.0%-5.5%) | 100.0% | 100.0% |

**Figure 1.**
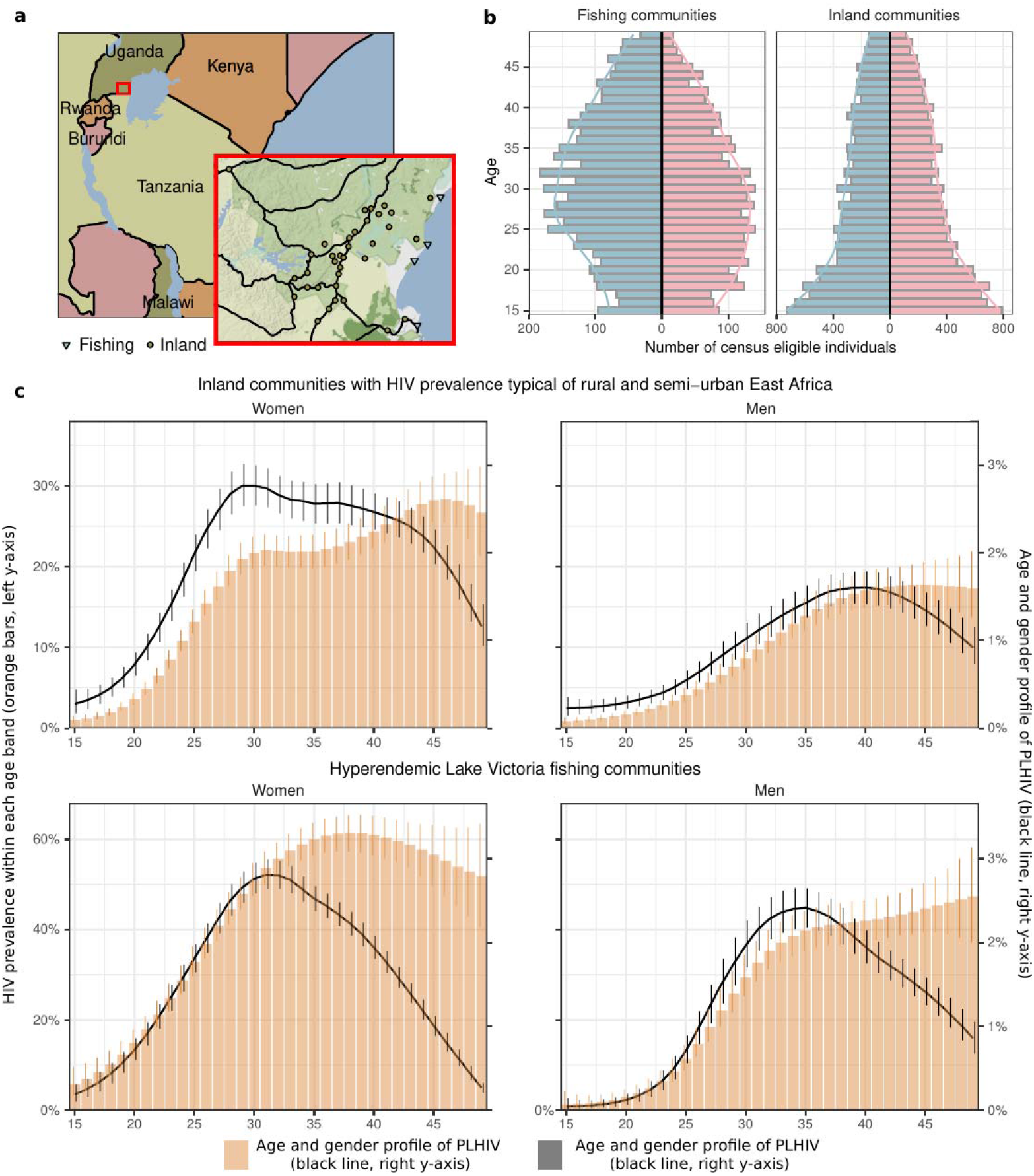
Age and gender profiles of HIV burden in the Rakai Community Cohort Study, June 2018-May 2019 (round 19). **(a)** Map of the fishing (triangles) and inland (circles) study communities within the Rakai region in south-central Uganda, produced using the ‘ggmap’ package version 3.0.2[28] in R (©Stadia Maps ©Stamen Design ©OpenMapTiles © OpenStreetMap contributor) **(b)** Population pyramids in fishing and inland communities. For each 1-year age band, the number of census-eligible individuals (colored bars) are shown for men(blue) and women (pink). Colored lines represent smoothed population size estimates. **(c)** The profile for the burden of HIV in the population is shown in black (line: posterior median estimate, linerange: 95% credible interval) and values shown on the right y-axis, for both community types (top, bottom) and both genders (left, right). For comparison, HIV seroprevalence within each 1-year age band is shown in orange (bars: posterior median estimates, error bar: 95% credible interval), and values shown on the left y-axis.

To describe the burden of HIV infection, we initially followed the typical approach to reporting HIV seroprevalence and estimated the proportion of individuals with HIV infection (i.e., positive HIV serostatus) in each 1-year age band (Figure 1c, orange). Despite close geographical proximity, HIV seroprevalence was higher in fishing communities along Lake Victoria than in the inland communities[19]. In 2019, HIV seroprevalence was 32.8% (95% posterior credible interval: 31.5%-34.2%) in fishing communities, versus 11.0% (10.5%-11.5%) in inland communities (Table 1), and it was larger among first-time participants (Supplementary Figure S3).

The distinct population pyramids of these communities also prompted us to characterize the HIV infection burden profile (Figure 1c). We found that this profile centered more strongly on younger populations than statistics of HIV seroprevalence within each 1-year age band might suggest, such that some younger populations with relatively lower HIV seroprevalence accounted for a higher fraction of the HIV infection burden. For example, by 2019, in inland communities half (51.4% (49.3%-53.5%)) of all women with HIV were 27-39 years old and half (53.1% (49.5%-56.3%)) of all men with HIV were 34-45 years old.

Differences in the gender burden of HIV infection across community types depended both on differences in seroprevalence and population structures. In 2019, HIV seroprevalence among women was 1.53 (1.40-1.66) fold higher than among men in fishing communities and 1.78 (1.61-1.98) fold higher among women than among men in inland communities. Further, as women accounted for a smaller proportion of the population in fishing communities (44.3%) as opposed to in inland communities (52.2%), women made up a smaller fraction of the PLHIV in fishing communities (55.2% (53.1%-57.3%)) rather than in inland communities (66.2% (63.8%-68.4%)).

### 3.2 Progress towards 95-95-95 targets

We next characterized progress towards “95-95-95” targets between 2013 and 2019 in the high-prevalence fishing communities that have since 2013 been prioritized for TasP, and in the inland communities where Universal Test and Treat was implemented beginning in late 2016. In total, 13,844 viral load measurements were taken in 6,677 of 6,705 (99.6%) survey participants with HIV over the four survey rounds. The number of people with viraemia decreased substantially from 2013 to 2019 (Table 2). Specifically, in inland communities, the proportion of women with HIV exhibiting viraemia decreased from 34.7% (32.3%-37.1%) in 2013 to 13.2% (11.5%-15.0%) in 2019, while the proportion of men with HIV exhibiting viraemia decreased from 50.7% (46.5%-54.8%) in 2019 to 26.3% (22.2%-30.8%) in 2013. In fishing communities, the proportion of women with HIV exhibiting viraemia decreased from 44.0% (40.9%-47.0%) in 2013 to 11.4% (9.3%-13.8%) in 2019, while the proportion of men with HIV exhibiting viraemia decreased from 64.7% (60.8%-68.5%) in 2013 to 18.1% (14.5%-22.3%) in 2019.

**Table 2.**
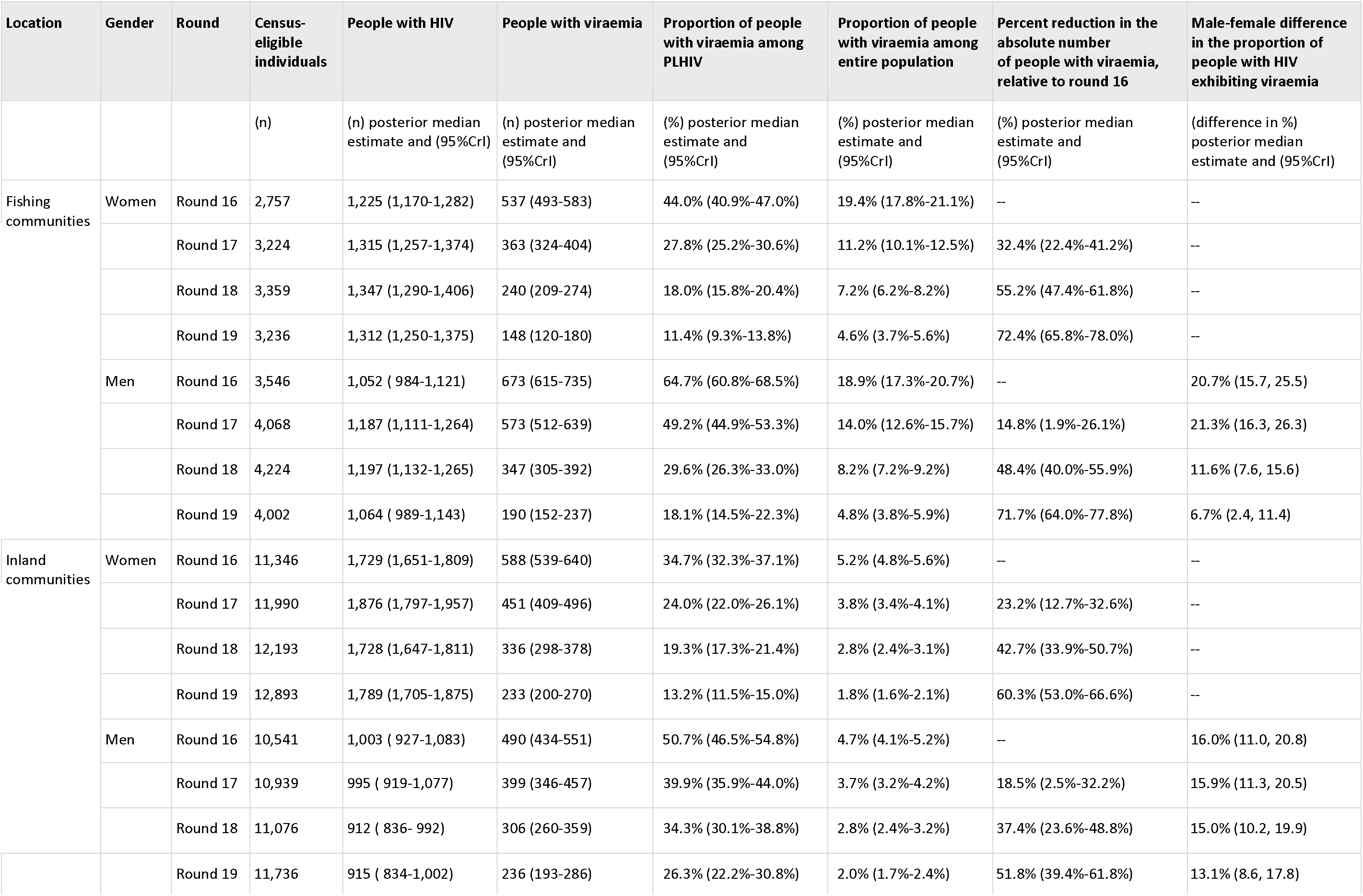
People exhibiting viraemia in the Rakai Community Cohort Study, from July 2013-January 2015 (round 16) to June 2018-May 2019 (round 19)

| Location | Gender | Round | Census-eligible individuals | People with HIV | People with viraemia | Proportion of people with viraemia among PLHIV | Proportion of people with viraemia among entire population | Percent reduction in the absolute number of people with viraemia, relative to round 16 | Male-female difference in the proportion of people with HIV exhibiting viraemia |
| --- | --- | --- | --- | --- | --- | --- | --- | --- | --- |
|  |  |  | (n) | (n) posterior median estimate and (95%CrI) | (n) posterior median estimate and (95%CrI) | (%) posterior median estimate and (95%CrI) | (%) posterior median estimate and (95%CrI) | (%) posterior median estimate and (95%CrI) | (difference in %) posterior median estimate and (95%CrI) |
| Fishing communities | Women | Round 16 | 2,757 | 1,225 (1,170-1,282) | 537 (493-583) | 44.0% (40.9%-47.0%) | 19.4% (17.8%-21.1%) | -- | -- |
|  |  | Round 17 | 3,224 | 1,315 (1,257-1,374) | 363 (324-404) | 27.8% (25.2%-30.6%) | 11.2% (10.1%-12.5%) | 32.4% (22.4%-41.2%) | -- |
|  |  | Round 18 | 3,359 | 1,347 (1,290-1,406) | 240 (209-274) | 18.0% (15.8%-20.4%) | 7.2% (6.2%-8.2%) | 55.2% (47.4%-61.8%) | -- |
|  |  | Round 19 | 3,236 | 1,312 (1,250-1,375) | 148 (120-180) | 11.4% (9.3%-13.8%) | 4.6% (3.7%-5.6%) | 72.4% (65.8%-78.0%) | -- |
|  | Men | Round 16 | 3,546 | 1,052 ( 984-1,121) | 673 (615-735) | 64.7% (60.8%-68.5%) | 18.9% (17.3%-20.7%) | -- | 20.7% (15.7, 25.5) |
|  |  | Round 17 | 4,068 | 1,187 (1,111-1,264) | 573 (512-639) | 49.2% (44.9%-53.3%) | 14.0% (12.6%-15.7%) | 14.8% (1.9%-26.1%) | 21.3% (16.3, 26.3) |
|  |  | Round 18 | 4,224 | 1,197 (1,132-1,265) | 347 (305-392) | 29.6% (26.3%-33.0%) | 8.2% (7.2%-9.2%) | 48.4% (40.0%-55.9%) | 11.6% (7.6, 15.6) |
|  |  | Round 19 | 4,002 | 1,064 ( 989-1,143) | 190 (152-237) | 18.1% (14.5%-22.3%) | 4.8% (3.8%-5.9%) | 71.7% (64.0%-77.8%) | 6.7% (2.4, 11.4) |
| Inland communities | Women | Round 16 | 11,346 | 1,729 (1,651-1,809) | 588 (539-640) | 34.7% (32.3%-37.1%) | 5.2% (4.8%-5.6%) | -- | -- |
|  |  | Round 17 | 11,990 | 1,876 (1,797-1,957) | 451 (409-496) | 24.0% (22.0%-26.1%) | 3.8% (3.4%-4.1%) | 23.2% (12.7%-32.6%) | -- |
|  |  | Round 18 | 12,193 | 1,728 (1,647-1,811) | 336 (298-378) | 19.3% (17.3%-21.4%) | 2.8% (2.4%-3.1%) | 42.7% (33.9%-50.7%) | -- |
|  |  | Round 19 | 12,893 | 1,789 (1,705-1,875) | 233 (200-270) | 13.2% (11.5%-15.0%) | 1.8% (1.6%-2.1%) | 60.3% (53.0%-66.6%) | -- |
|  | Men | Round 16 | 10,541 | 1,003 ( 927-1,083) | 490 (434-551) | 50.7% (46.5%-54.8%) | 4.7% (4.1%-5.2%) | -- | 16.0% (11.0, 20.8) |
|  |  | Round 17 | 10,939 | 995 ( 919-1,077) | 399 (346-457) | 39.9% (35.9%-44.0%) | 3.7% (3.2%-4.2%) | 18.5% (2.5%-32.2%) | 15.9% (11.3, 20.5) |
|  |  | Round 18 | 11,076 | 912 ( 836- 992) | 306 (260-359) | 34.3% (30.1%-38.8%) | 2.8% (2.4%-3.2%) | 37.4% (23.6%-48.8%) | 15.0% (10.2, 19.9) |
|  |  | Round 19 | 11,736 | 915 ( 834-1,002) | 236 (193-286) | 26.3% (22.2%-30.8%) | 2.0% (1.7%-2.4%) | 51.8% (39.4%-61.8%) | 13.1% (8.6, 17.8) |

Considering the number of people exhibiting viraemia in the entire population, by 2019, there were 233 (200-270) women and 236 (193-286) men with viraemia in inland communities, corresponding to 1.8% (1.6%-2.1%) and 2.0% (1.7%-2.4%) of the census-eligible female and male populations, respectively. In fishing communities, there were 148 (120-180) women and 190 (152-237) men exhibiting viraemia in 2019, corresponding to 4.6% (3.7%-5.6%) and 4.8% (3.8%-5.9%) of census-eligible women and men, respectively (Tables 1, 2). Thus, while suppression levels among PLHIV were higher in fishing communities compared to inland communities in both men and women, suppression levels in the population, i.e. among census-eligible individuals, were lower in fishing communities rather than inland communities due to the overall higher HIV prevalence.

### 3.3 Age and gender profiles of the burden of HIV viraemia

To characterize the age and gender-specific populations in which viraemia concentrates, we first considered the proportion of PLHIV exhibiting viraemia stratified by age and gender. By 2019, in inland communities, the proportion of PLHIV exhibiting viraemia was highest in men aged 23-27 years (49.8% (39.9%-60.3%)) (Figure 2), showing also that conventional 5-year age banding (15-19 years, 20-24 years …) can mask the actual age groups in which prevalence of viraemia peaks. In fishing communities, the proportion of PLHIV exhibiting viraemia in 2019 was highest in men aged 28-32 years (28.2% (20.0%-41.2%)) (Figure 2). Across all age groups, the proportion of PLHIV exhibiting viraemia was larger in first-time participants (Supplementary Figures S4-5), and lower, or equal in women than men. By 2019, among women, this proportion was highest in women aged 15-17 years in inland communities (30.3% (18.7%-44.0%)) and in women aged 18-22 years in fishing communities (23.9% (16.2%-34.0%)).

**Figure 2:**
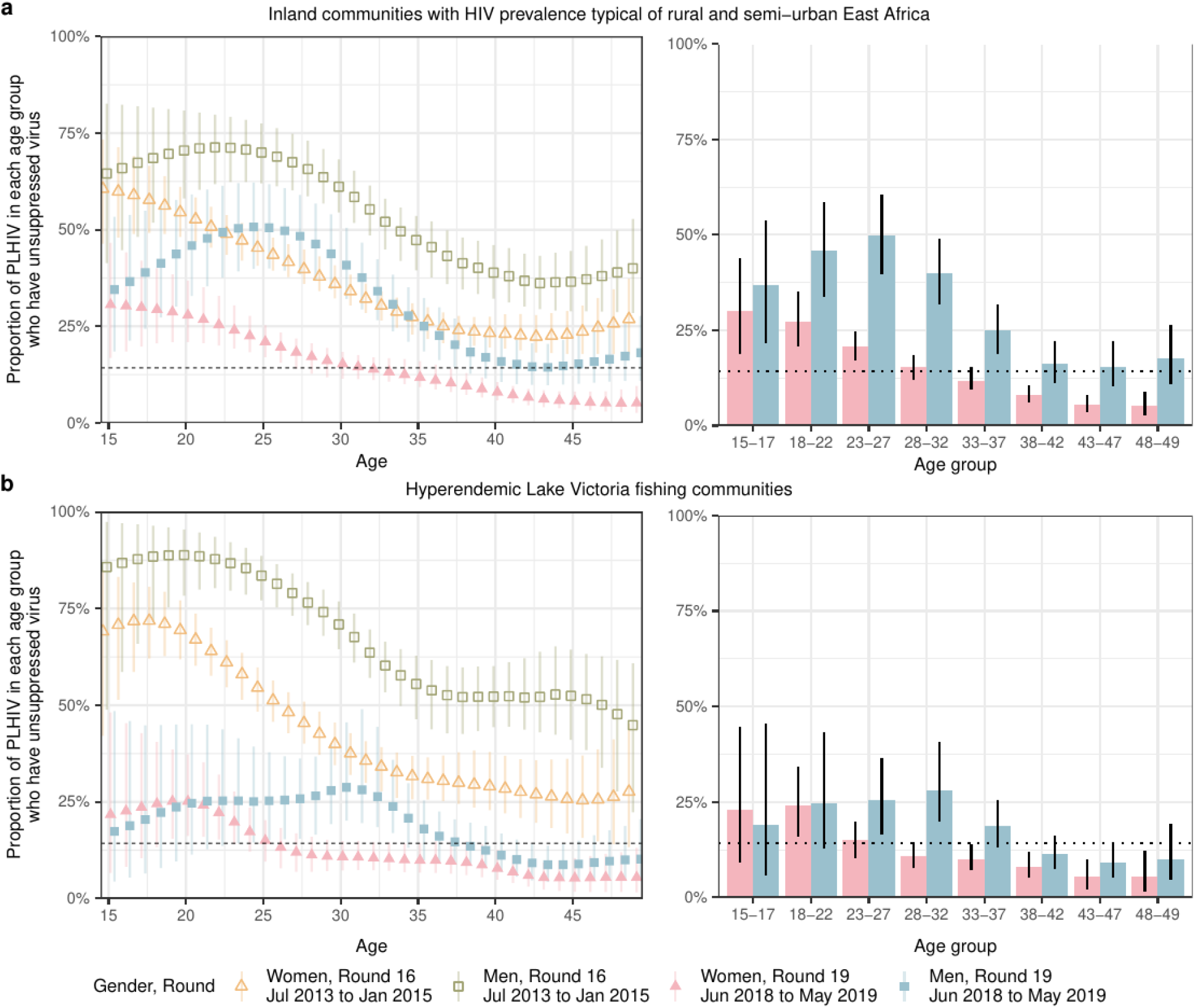
Trends in the proportions of PLHIV who have unsuppressed viraemia in the Rakai Community Cohort Study. **(a)** Estimated proportions of people with HIV in 1-year age bands who have unsuppressed viraemia, in men and women of inland communities. On the left, time trends in the proportion of PLHIV with unsuppressed viraemia from round 16 (July 2013-January 2015) to round 19 (June 2018 to May 2019) are shown for men and women by 1-year age bands (points: posterior median estimates, linerange: 95% CrIs). On the right, posterior median estimates in the proportion of PLHIV with unsuppressed viraemia in round 19 are shown for men and women by 5-year age bands (bars: posterior median estimates, linerange: 95% CrIs). The horizontal dotted line represents the 14% suppression threshold implied by 95-95-95 targets. **(b)** Same for PLHIV in fishing communities.

Then, we accounted for the distinct population structure in inland and fishing communities to characterize the population subgroups in which viraemia is concentrating. Figure 3a compares the age and gender specific prevalence of viraemia among PLHIV to the viraemia burden profile in the population. We found that viraemia did not center in the age groups currently furthest from achieving “95-95-95” targets. Specifically, in inland communities, women aged 15-19 years were furthest from achieving the “95-95-95” targets in 2019, but we estimate they contributed only 4.9% (3.0%-7.3%) to people exhibiting viraemia. Instead, women aged 20-24 years and 25-29 years each contributed approximately 10% to people exhibiting viraemia in 2019, although these age groups were either close to or had already exceeded the “95-95-95” targets. Similarly, in inland communities, men aged 20-24 years were furthest from achieving “95-95-95” targets in 2019, but we estimate they contributed only 5.4% (3.9%-7.4%) to the burden of viraemia. Rather, men aged 25-29 years and 30-34 years each contributed approximately 10% to the burden of viraemia in 2019. Similar patterns were observed in fishing communities. In 2019, women aged 15-19 were one of the few groups that had not yet met “95-95-95” targets, but this group only contributed 5.1% (2.5%-7.8%) to the burden of viraemia. Instead, women aged 30-34 had met the targets but contributed 8.9% (6.7%-12.6%) to the burden of viraemia. Among men, age groups from 20 to 34 years exhibited similar levels of viral suppression among PLHIV. However, there were more men aged 30-34 exhibiting viraemia as compared to men aged 20-24. The former amounted to 19.6% (14.8%-25.9%) of the burden of viraemia as compared to 3.0% (1.6%-5.2%) in the latter group.

**Figure 3.**
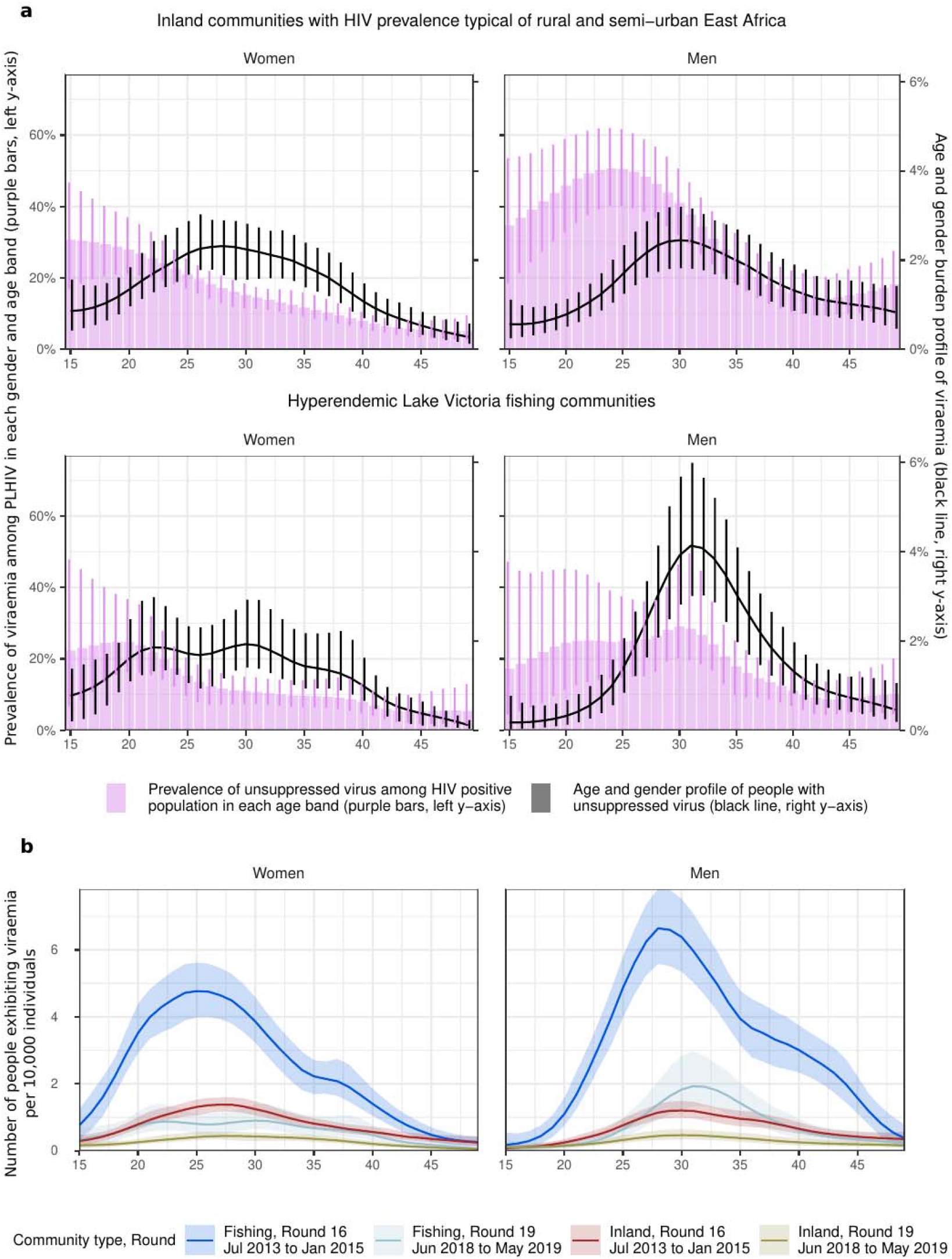
Age and gender metrics for people exhibiting viraemia. **(a)** Estimates for the prevalence of viraemia among PLHIV (median: purple bars, lines: 95% CrI, right y-axis) in each gender and age band are compared to the burden profile of viraemia (median: black curve, lines: 95% CrI, left y-axis) for inland (top row) and fishing communities (bottom row). **(b)** Time trends in the burden profiles of viraemia for both men and women (facets), colored by community type (inland or fishing) and survey round (16 or 19) colors (line: posterior median estimate, ribbon: 95% credible interval). The profiles were multiplied by the population prevalence of viral suppression to visualize shifts in the size of the population exhibiting viraemia.

Figure 3b depicts temporal changes in absolute suppression levels, as well as shifts in the profile of the burden of viraemia. Even though we found a steady increase in the median age of PLHIV by gender and community strata, the age profile of people exhibiting viraemia remained relatively stable over time (Supplementary Table S3).

## 4. Discussion

This study estimated the age and gender burden profiles of HIV infection and HIV viraemia in south-central Uganda using data from communities with HIV risk and demographic profiles typical of rural and semi-urban ESA and data from Lake Victoria fishing communities with high HIV seroprevalence and atypical demographic profiles. Overall, the burden profiles revealed important differences by geographic location, gender and age group. Following the earlier expansion of TasP programs, fishing communities witnessed faster increases in suppression levels among PLHIV than in inland communities, with overall levels surpassing the “95-95-95” targets by 2019. However, the burden of HIV viraemia remained larger in fishing communities, where the proportion of individuals exhibiting viraemia was approximately 2.5 times larger than in inland communities, reflecting higher HIV prevalence in fishing communities. When analyzing the burden of viraemia by gender, we found that in both community types, most individuals exhibiting viraemia were men, although fewer men have HIV than women. When also considering age, we found that the age groups that contributed most to the burden of viraemia were not necessarily the ones that were furthest from achieving the “95-95-95” targets. For example, in inland communities, individuals aged 25-34 contributed to approximately 40% of the burden of viraemia but had higher suppression levels than younger age groups.

Overall, our results show that the metrics considered by the “95-95-95” targets are insufficient to characterize the population exhibiting viraemia, and hence at risk of transmitting HIV. For example, by 2019, fishing communities had an overall prevalence of suppression among PLHIV that surpassed “95-95-95” targets, while inland communities had not yet achieved them. However, individuals exhibiting viraemia represented a larger proportion of the population in fishing communities as opposed to inland communities, consistent with HIV incidence remaining higher in the former communities despite additional interventions such as PrEP[29]. This is largely explained by differences in HIV prevalence and implies that reaching “95-95-95” by 2025 may be insufficient to lower incidence below elimination threshold[30], especially in countries with elevated burden of HIV[31].

Moreover, since 2020 the updated UNAIDS targets recommend the “95-95-95” goals to be met within subpopulations stratified by age and gender[32]. In practice, this supports the prioritization of interventions to those subgroups that are furthest from achieving the targets and improving barriers and inequalities in treatment coverage[18]. However, these subgroups may not disproportionately contribute to the burden of HIV viraemia. For example, in inland communities, we found that young individuals aged 15-24 constituted approximately two fifths of the census-eligible population, but only one fifth of the population exhibiting viremia. In fact, the lower suppression levels commonly reported among young PLHIV[12,33–40] are counterbalanced by the small absolute number of young PLHIV. This observation is typically missing from studies supporting prioritization of younger age-groups in ESA based on the UNAIDS targets^45^. However, there are instances in which higher suppression levels and HIV seroprevalence lead to balanced profiles. For instance, although most PLHIV are women, there are similar numbers of men and women exhibiting viraemia in inland communities. Taken together, these findings support the need to reach and treat a wide subset of the population: men aged 25-34 and women aged 20-39, who disproportionately contribute to unsuppressed viraemia.

The longitudinal nature of the study further allowed us to analyze temporal dynamics in the age profile for the burden of HIV. From 2013 to 2019, we found an increase in the median age of PLHIV of at least 2 years across community types and genders. Although incidence and HIV mortality are decreasing, a shift in this direction is not guaranteed. In inland communities, younger individuals constitute an increasing proportion of the population, and therefore small reductions in incidence may still be consistent with increasing numbers of PLHIV in these age groups. More generally, the population is expected to grow at an exponential rate in Sub-Saharan Africa[41], implying that stagnating incidence rates may potentially shift the age profile to younger age groups. Continued efforts to track the changing demographics of countries in the region, for example through population censuses, would provide helpful in planning long-term HIV interventions, as well as generalizing to other Sustainable Development Goals[42].

While we assessed both age and gender burden profiles of HIV infection and HIV viraemia in this study, the latter metric provides a cross-sectional description of individuals at most at risk of HIV mortality and onward transmission. Hence, we propose the viraemia burden profile as a new metric for HIV control. Critically we observed that the burden profiles of viraemia revealed similar relative contributions of men and women, and larger relative contributions of middle-aged individuals (25-34) than younger individuals (15-24). We further show that these important observations are obscured by age-gender stratified “95-95-95” targets. Furthermore, unlike the “95-95-95” targets[14,15], population burden of HIV viraemia has been shown to be strongly predictive of population HIV incidence[17,43].

This study has several limitations. First, due to the RCCS definition of census-eligible individuals in the study period, our study population is limited to individuals aged 15-49 years. Accordingly, we are not able to quantify the growing seroprevalence of HIV among individuals aged 50 or more, nor viral suppression outcomes in children (<15 years) living with HIV. Importantly, the reported age and gender specific percentages in the burden profiles in the census-eligible population aged 15-49 years should not be interpreted as fractions of the entire population, as they would be overestimates. Second, our study only considered data from Uganda, and our results may not directly generalize to other settings. However epidemiological patterns reported here are typical of ESA, such as: expansive population pyramids[27], patterns in HIV prevalence by age and gender[44], and gender gaps in suppression levels[12,33–40]. These observations suggests that the major contributions of individuals aged 25-34 to the burden of viraemia may generalize across the subcontinent. Third, to generalize our results to the entire census-eligible population, we assumed that first-time study participants were representative of non-participants in terms of age and gender-specific prevalence and viral suppression levels. Despite similar HIV seroprevalence estimates, our estimates of viral suppression among first-time participants were lower than for all participants. Generalizing the results to the census-eligible population avoided overestimation of viral suppression levels and allowed us to match our analyses’ denominators to the ones defined in the UNAIDS goals. Fourth, the age profile of the population exhibiting viraemia may not match the age profile of the sources of HIV infections, but rather the potential sources of HIV transmission. The difference between the two populations can be explained by heterogeneities in transmission risk among individuals exhibiting viraemia. For example, phylogenetic studies in Uganda[21], Zambia[45], and Botswana[46] have estimated highest transmission risks in men aged 30-34 and women aged 20-24, and much lower for example among individuals aged more than 50. These results hence reinforce the importance of the subpopulations contributing most strongly to the burden of viraemia described in this paper.

## 5. Conclusions

In summary, prioritizing HIV prevention interventions across age and gender subgroups solely based on the “95-95-95” targets may not be the most effective way to reduce the number of people exhibiting viraemia, and hence at risk of HIV mortality and transmitting HIV. Importantly, these metrics ignore underlying population structures and differences in HIV seroprevalence by subgroups, which are integral to the calculation of the age and gender profile for the burden of HIV viraemia. Future surveillance efforts should prioritize population censuses and population-level measures of HIV viraemia to better direct HIV control and elimination efforts, particularly in settings at or achieving the “95-95-95” targets.

## Competing interests

MKG, LCW report grants from the National Institutes of Health during the conduct of this study. ABr, MM report an EPSRC PhD studentship during the conduct of this study. OR acknowledges grants from the Bill and Melinda Gates Foundation (OPP1175094 to C.Fraser., OPP1084362 to D. Pillay), the Engineering and Physical Sciences Research Council (EP/X038440/1), and the Moderna Charitable Foundation during the conduct of this study. This arrangement has been reviewed and approved by the Johns Hopkins University in accordance with its conflict-of-interest policies.

## Authors’ contributions

OR, MKG, SJR, RS designed the study. OR, MKG, SJR, JK oversaw the study. RG, RS, GK, OL, TQ, FN, LWC, SJR, JK, GN, MKG oversaw and performed data collection. ABr, IK, ABl, MM, MKG, OR contributed to the analysis. ABr, MKG, OR wrote the first draft. All authors discussed the results and contributed to the revision of the final manuscript.

## Author information

[Optional]

## Acknowledgements

We thank all contributors, program staff and participants to the Rakai Community Cohort Study; all members of the PANGEA-HIV consortium, the Rakai Health Sciences Program; the Imperial College Research Computing Service (https://doi.org/10.14469/hpc/2232) the Office of Cyberinfrastructure and Computational Biology at the National Institute for Allergy and Infectious Diseases for data management support, and Zulip for sponsoring team communications through the Zulip Cloud Standard chat app.

## Funding

This study was supported by the National Institute of Allergy and Infectious Diseases [U01AI075115 to RHG, R01AI087409 to RHG, U01AI100031 to RHG, ZIAAI001040 to TCQ, R01AI43333 to LWC]; the National Institute of Mental Health [F31MH095649 to Dr Jennifer Wagman, R01MH099733 to Ned Sacktor and MJW, R01MH107275 to LWC]; the Division of Intramural Research of the National Institute for Allergy and Infectious Diseases [TCQ, OL, SJR], NIAID [K01AA024068 to Dr Jennifer Wagman]; the Johns Hopkins University Center for AIDS Research [2P30AI094189 to Dr Richard Chaisson]; the U.S. President’s Emergency Plan for AIDS Relief (PEPFAR) through the Centers for Disease Control and Prevention [NU2GGH000817 to RHSP]; the Engineering and Physical Sciences Research Council through the EPSRC Centre for Doctoral Training in Modern Statistics and Statistical Machine Learning at Imperial and Oxford [EP/S023151/1 to Prof Axel Gandy]. The funders had no role in study design, data collection and analysis, decision to publish or preparation of the manuscript. The findings and conclusions in this report are those of the author(s) and do not necessarily represent the official position of the Centers for Disease Control and Prevention.

## Disclaimer

For the purpose of open access, the author has applied a ‘Creative Commons Attribution’ (CCBY) license to any Author Accepted Manuscript version arising.

## Data Availability Statement

Data necessary to reproduce the analyses are available on Zenodo (https://doi.org/10.5281/zenodo.10955672) as open access dataset under the CC-BY-4.0 license. Code to reproduce all analyses is freely available on GitHub version 1.1.2 under the CC-BY-4.0 license at the repository (https://github.com/MLGlobalHealth/longi_viral_loads).

## Supporting Information

[Optional]

Supporting Information file 1: Title of Supporting Information file

Information on file format. Brief description of file content.

## List of abbreviations

ART: Antiretroviral Therapy
HIV: Human Immunodeficiency Virus
PLHIV: People living with HIV
RCCS: Rakai Community Cohort Study
RHSP: Rakai Health Sciences Program
UNAIDS: Joint United Nations Programme on HIV/AIDS
WHO: World Health Organization

## References

1 UNAIS. The path that ends AIDS: UNAIDS Global AIDS Update 2023. 2023.https://www.unaids.org/en/resources/documents/2023/global-aids-update-2023

2 Mills EJ, Bakanda C, Birungi J, Chan K, Ford N, Cooper CL, et al. Life Expectancy of Persons Receiving Combination Antiretroviral Therapy in Low-Income Countries: A Cohort Analysis From Uganda. Ann Intern Med 2011; 155:209.

3 Herbst AJ, Cooke GS, Bärnighausen T, KanyKany A, Tanser F, Newell M. Adult mortality and antiretroviral treatment roll-out in rural KwaZulu-Natal, South Africa. Bull World Health Org 2009; 87:754–762.

4 Reniers G, Slaymaker E, Nakiyingi-Miiro J, Nyamukapa C, Crampin AC, Herbst K, et al. Mortality trends in the era of antiretroviral therapy: evidence from the Network for Analysing Longitudinal Population based HIV/AIDS data on Africa (ALPHA). AIDS 2014; 28:S533–S542.

5 Attia S, Egger M, Müller M, Zwahlen M, Low N. Sexual transmission of HIV according to viral load and antiretroviral therapy: systematic review and meta-analysis. AIDS 2009; 23:1397–1404.

6 Cohen MS, Chen YQ, McCauley M, Gamble T, Hosseinipour MC, Kumarasamy N, et al. Prevention of HIV-1 Infection with Early Antiretroviral Therapy. N Engl J Med 2011; 365:493–505.

7 Stover J, Glaubius R, Teng Y, Kelly S, Brown T, Hallett TB, et al. Modeling the epidemiological impact of the UNAIDS 2025 targets to end AIDS as a public health threat by 2030. PLoS Med 2021; 18:e1003831.

8 Godfrey-Faussett P, Frescura L, Abdool Karim Q, Clayton M, Ghys PD, (on behalf of the 2025 prevention targets working group). HIV prevention for the next decade: Appropriate, person-centred, prioritised, effective, combination prevention. PLoS Med 2022; 19:e1004102.

9 UNAIDS. Countries. https://www.unaids.org/en/regionscountries/countries (accessed 1 Nov2022).

10 United Nations. Sustainable Deveopment Goals. United Nations Sustainable Development. https://www.un.org/sustainabledevelopment/sustainable-development-goals/ (accessed 13 Mar2024).

11 Sachs JD. From Millennium Development Goals to Sustainable Development Goals. The Lancet 2012; 379:2206–2211.

12 Mine M, Stafford KA, Laws RL, Marima R, Lekone P, Ramaabya D, et al. Progress towards the UNAIDS 95-95-95 targets in the Fifth Botswana AIDS Impact Survey (BAIS V 2021): a nationally representative survey. The Lancet HIV 2024; 0. doi:10.1016/S2352-3018(24)00003-1

13 Maheu-Giroux M, Mishra S. Evidence with 95-95-95 that ambitious is feasible. The Lancet HIV 2024; 0. doi:10.1016/S2352-3018(24)00028-6

14 Miller WC, Powers KA, Smith MK, Cohen MS. Community viral load as a measure for assessment of HIV treatment as prevention. The Lancet Infectious Diseases 2013; 13:459–464.

15 Solomon SS, Mehta SH, McFall AM, Srikrishnan AK, Saravanan S, Laeyendecker O, et al. Community viral load, antiretroviral therapy coverage, and HIV incidence in India: a cross-sectional, comparative study. The Lancet HIV 2016; 3:e183–e190.

16 Tanser F, Vandormael A, Cuadros D, Phillips AN, De Oliveira T, Tomita A, et al. Effect of population viral load on prospective HIV incidence in a hyperendemic rural African community. Sci Transl Med 2017; 9:eaam8012.

17 Larmarange J, Bachanas P, Skalland T, Balzer LB, Iwuji C, Floyd S, et al. Population-level viremia predicts HIV incidence at the community level across the Universal Testing and Treatment Trials in eastern and southern Africa. PLOS Glob Public Health 2023; 3:e0002157.

18 Frescura L, Godfrey-Faussett P, Feizzadeh A. A, El-Sadr W, Syarif O, Ghys PD, et al. Achieving the 95 95 95 targets for all: A pathway to ending AIDS. PLoS ONE 2022; 17:e0272405.

19 Chang LW, Grabowski MK, Ssekubugu R, Nalugoda F, Kigozi G, Nantume B, et al. Heterogeneity of the HIV epidemic in agrarian, trading, and fishing communities in Rakai, Uganda: an observational epidemiological study. The Lancet HIV 2016; 3:e388–e396.

20 Grabowski MK, Serwadda DM, Gray RH, Nakigozi G, Kigozi G, Kagaayi J, et al. HIV Prevention Efforts and Incidence of HIV in Uganda. N Engl J Med 2017; 377:2154–2166.

21 Monod M, Brizzi A, Galiwango RM, Ssekubugu R, Chen Y, Xi X, et al. Longitudinal population-level HIV epidemiological and genomic surveillance highlights growing gender disparity of HIV transmission in Uganda. Epidemiology; 2023. doi:10.1101/2023.03.16.23287351

22 Galiwango RM, Musoke R, Lubyayi L, Ssekubugu R, Kalibbala S, Ssekweyama V, et al. Evaluation of current rapid HIV test algorithms in Rakai, Uganda. Journal of Virological Methods 2013; 192:25–27.

23 Ssempijja V, Chang LW, Nakigozi G, Ndyanabo A, Quinn TC, Cobelens F, et al. Results of Early Virologic Monitoring May Facilitate Differentiated Care Monitoring Strategies for Clients on ART, Rakai, Uganda. Open Forum Infectious Diseases 2018; 5:ofy212.

24 Ssempijja V, Nason M, Nakigozi G, Ndyanabo A, Gray R, Wawer M, et al. Adaptive Viral Load Monitoring Frequency to Facilitate Differentiated Care: A Modeling Study From Rakai, Uganda. Clin Infect Dis 2020; 71:1017–1021.

25 Grabowski MK, Patel EU, Nakigozi G, Ssempijja V, Ssekubugu R, Ssekasanvu J, et al. Prevalence and Predictors of Persistent Human Immunodeficiency Virus Viremia and Viral Rebound After Universal Test and Treat: A Population-Based Study. The Journal of Infectious Diseases 2021; 223:1150–1160.

26 Consolidated Guidelines on the Use of Antiretroviral Drugs for Treating and Preventing HIV Infection: Recommendations for a Public Health Approach. 2nd ed. Geneva: World Health Organization; 2016. http://www.ncbi.nlm.nih.gov/books/NBK374294/ (accessed 20 Oct2023).

27 Population Pyramids of the World from 1950 to 2100. PopulationPyramid.net. https://www.populationpyramid.net/ uganda/2020/ (accessed 9 Aug2023).

28 Kahle D, Wickham H. ggmap: Spatial Visualization with ggplot2. The R Journal 2013; 5:144–161.

29 Ssempijja V, Nakigozi G, Ssekubugu R, Kagaayi J, Kigozi G, Nalugoda F, et al. High Rates of Pre-exposure Prophylaxis Eligibility and Associated HIV Incidence in a Population With a Generalized HIV Epidemic in Rakai, Uganda. JAIDS Journal of Acquired Immune Deficiency Syndromes 2022; 90:291–299.

30 Granich RM, Gilks CF, Dye C, De Cock KM, Williams BG. Universal voluntary HIV testing with immediate antiretroviral therapy as a strategy for elimination of HIV transmission: a mathematical model. The Lancet 2009; 373:48–57.

31 Dimitrov D, Moore JR, Deborah J. Donnell, Marie-Claude Boily. Achieving 95-95-95 may not be enough to end the AIDS epidemic in South Africa. 2020.croiconference.org/wp-content/uploads/sites/2/posters/2020/1430_6_Dimitrov_01095.pdf (accessed 11 Mar2024).

32 UNAIDS. Understanding Fast-Track: accelerating actoin to end the AIDS epidemic by 2030.; 2015. https://www.unaids.org/sites/default/files/media_asset/201506_JC2743_Understanding_FastTrack_en.pdf

33 Lesotho Summary Sheet 2020. https://phia.icap.columbia.edu/wp-content/uploads/2021/08/53059_14_LePHIA_Summary-sheet_with-coat-of-arms_WEB_v2.pdf

34 Eswatini Summary Sheet 2021. https://phia.icap.columbia.edu/wp-content/uploads/2022/12/53059_14_SHIMS3_Summary-sheet-Web.pdf

35 Malawi Summary Sheet 2020-2021. https://phia.icap.columbia.edu/wp-content/uploads/2022/03/110322_MPHIA_Summary-sheet-English.pdf

36 Mozambique Summary Sheet 2021. https://phia.icap.columbia.edu/wp-content/uploads/2022/12/53059_14_INSIDA_Summary-sheet-Web.pdf

37 Uganda Summary Sheet 2020-2021. https://phia.icap.columbia.edu/wp-content/uploads/2022/08/UPHIA-Summary-Sheet-2020.pdf

38 Pathmanathan I, Nelson R, De Louvado A, Thompson R, Pals S, Casavant I, et al. High Coverage of Antiretroviral Treatment With Annual Home-Based HIV Testing, Follow-up Linkage Services, and Implementation of Test and Start: Findings From the Chókwè Health Demographic Surveillance System, Mozambique, 2014–2019. JAIDS Journal of Acquired Immune Deficiency Syndromes 2021; 86:e97–e105.

39 Pillay T, Cornell M, Fox MP, Euvrard J, Fatti G, Technau K-G, et al. Recording of HIV Viral Loads and Viral Suppression in South African Patients Receiving Antiretroviral Treatment: A Multicentre Cohort Study. Antiviral Therapy 2020; 25:257–266.

40 Nyaboke R, Ramadhani HO, Lascko T, Awuor P, Kirui E, Koech E, et al. Factors associated with adherence and viral suppression among patients on second-line antiretroviral therapy in an urban HIV program in Kenya. SAGE Open Medicine 2023; 11:205031212311623.

41 Musa SS, Zhao S, Wang MH, Habib AG, Mustapha UT, He D. Estimation of exponential growth rate and basic reproduction number of the coronavirus disease 2019 (COVID-19) in Africa. Infect Dis Poverty 2020; 9:96.

42 Sankoh O, on behalf of the INDEPTH Network and partners. Why population-based data are crucial to achieving the Sustainable Development Goals. International Journal of Epidemiology 2017; 46:4– 7.

43 Patel EU, Solomon SS, Lucas GM, McFall AM, Srikrishnan AK, Kumar MS, et al. Temporal change in population-level prevalence of detectable HIV viraemia and its association with HIV incidence in key populations in India: a serial cross-sectional study. The Lancet HIV 2021; 8:e544–e553.

44 Dwyer-Lindgren L, Cork MA, Sligar A, Steuben KM, Wilson KF, Provost NR, et al. Mapping HIV prevalence in sub-Saharan Africa between 2000 and 2017. Nature 2019; 570:189–193.

45 Hall M, Golubchik T, Bonsall D, Abeler-Dörner L, Limbada M, Kosloff B, et al. Demographics of people who transmit HIV-1 in Zambia: a molecular epidemiology analysis in the HPTN-071 PopART study. HIV/AIDS; 2021. doi:10.1101/2021.10.04.21263560

46 Magosi LE, Zhang Y, Golubchik T, DeGruttola V, Tchetgen Tchetgen E, Novitsky V, et al. Deep-sequence phylogenetics to quantify patterns of HIV transmission in the context of a universal testing and treatment trial – BCPP/Ya Tsie trial. eLife 2022; 11:e72657.

