## Supplementary Materials for "Age and gender profiles of HIV infection burden and viraemia: novel metrics for HIV epidemic control in African populations with high antiretroviral therapy coverage"

A. Brizzi et al.

#### List of Tables

S1 Census eligible population size and participation rates . . . S2
S2 Characteristics of RCCS first-time participants by community type, gender and survey round, 2013-2019. . . . S3
S3 Age profile of individuals with HIV and individuals with unsuppressed virus in the population by community type, gender and survey round, 2013-2019 . . . S4
S4 Convergence diagnostics for all models . . . S5

#### List of Figures

S1 Data flowchart. . . . S7
S2 RCCS population size estimates and participation rates, 2013-2019 . . . S8
S3 Age- and gender-specific HIV prevalence estimates among study participants and first-time participants, 2013-2019. . . . S9
S4 Age- and gender-specific viral load suppression rate estimates among study participants and first-time participants with HIV, 2013-2019. . . . S10
S5 Age- and gender-specific viral load suppression rate estimates of among census-eligible people with HIV. . . . S11
S6 Consistency of fitted models with available data. . . . S12

#### Contents

**S1 Extended Methods** **S13**
S1.1 Population size and participation rates . . . S13
S1.2 Population-level HIV prevalence estimates by age and gender . . . S13
S1.3 Age and gender profiles of people living with HIV . . . S14
S1.4 Population-level prevalence of viraemia estimates by age and gender among PLHIV . S14
S1.5 Age and gender profile of people exhibiting viraemia . . . S15
S1.6 Age quantiles for burden profiles by gender . . . S15

31 # Supplementary Tables

| Location | Gender | Survey round | Number of census eligible individuals | Number of participants | Participation rates |
|---|---|---|---|---|---|
| Fishing communities | Women | Round 16 07/13-01/15 | 2,757 | 1,867 | 67.7% |
| | | Round 17 02/15-09/16 | 3,224 | 2,077 | 64.4% |
| | | Round 18 10/16-05/18 | 3,359 | 2,225 | 66.2% |
| | | Round 19 06/18-05/19 | 3,236 | 1,968 | 60.8% |
| | Men | Round 16 07/13-01/15 | 3,546 | 2,067 | 58.3% |
| | | Round 17 02/15-09/16 | 4,068 | 2,163 | 53.2% |
| | | Round 18 10/16-05/18 | 4,224 | 2,535 | 60% |
| | | Round 19 06/18-05/19 | 4,002 | 2,123 | 53% |
| Inland communities | Women | Round 16 07/13-01/15 | 11,346 | 7,816 | 68.9% |
| | | Round 17 02/15-09/16 | 11,990 | 8,377 | 69.9% |
| | | Round 18 10/16-05/18 | 12,193 | 8,331 | 68.3% |
| | | Round 19 06/18-05/19 | 12,893 | 8,495 | 65.9% |
| | Men | Round 16 07/13-01/15 | 10,541 | 6,256 | 59.3% |
| | | Round 17 02/15-09/16 | 10,939 | 6,716 | 61.4% |
| | | Round 18 10/16-05/18 | 11,076 | 6,722 | 60.7% |
| | | Round 19 06/18-05/19 | 11,736 | 6,765 | 57.6% |

Supplementary Table S1: **Census eligible population size and participation rates by community type, gender and survey round.**

| Location | Gender | Survey round | First-time participants (n) | First-time participants with HIV (n) | First-time participants exhibiting viraemia (n) | Ratio of first-time participants with HIV relative to round 16 | Ratio of first-time participants exhibiting viraemia relative to round 16 |
|---|---|---|---|---|---|---|---|
| Fishing communities | Women | Round 16 07/13-01/15 | 492 | 198 | 100 | 1.00 | 1.00 |
| | | Round 17 02/15-09/16 | 670 | 241 | 87 | 1.22 | 0.87 |
| | | Round 18 10/16-05/18 | 688 | 231 | 57 | 1.17 | 0.57 |
| | | Round 19 06/18-05/19 | 540 | 176 | 29 | 0.89 | 0.29 |
| | Men | Round 16 07/13-01/15 | 529 | 122 | 88 | 1.00 | 1.00 |
| | | Round 17 02/15-09/16 | 577 | 134 | 77 | 1.1 | 0.88 |
| | | Round 18 10/16-05/18 | 766 | 159 | 57 | 1.3 | 0.65 |
| | | Round 19 06/18-05/19 | 505 | 103 | 21 | 0.84 | 0.24 |
| Inland communities | Women | Round 16 07/13-01/15 | 2,553 | 310 | 131 | 1.00 | 1.00 |
| | | Round 17 02/15-09/16 | 2,421 | 324 | 102 | 1.05 | 0.78 |
| | | Round 18 10/16-05/18 | 2,318 | 235 | 76 | 0.76 | 0.58 |
| | | Round 19 06/18-05/19 | 2,447 | 238 | 43 | 0.77 | 0.33 |
| | Men | Round 16 07/13-01/15 | 2,028 | 118 | 67 | 1.00 | 1.00 |
| | | Round 17 02/15-09/16 | 1,898 | 108 | 58 | 0.92 | 0.87 |
| | | Round 18 10/16-05/18 | 1,846 | 92 | 43 | 0.78 | 0.64 |
| | | Round 19 06/18-05/19 | 1,937 | 91 | 30 | 0.77 | 0.45 |

Supplementary Table S2: **Characteristics of RCCS first-time participants by community type, gender and survey round, 2013-2019.**

| Community type | Gender | Round | Age profile of individuals with HIV | | | Age profile of individuals with unsuppressed virus | | |
|---|---|---|---|---|---|---|---|---|
| | | | 25% quantile | Median | 75% quantile | 25% quantile | Median | 75% quantile |
| Fishing | Female | Round 16 | 24.4 | 29.3 | 34.9 | 22.2 | 26.7 | 32.2 |
| | | 07/13-01/15 | (24.2-24.5) | (29.2-29.5) | (34.7-35.0) | (22.0-22.4) | (26.5-27.0) | (31.9-32.6) |
| | | Round 17 | 24.5 | 29.3 | 35.0 | 23.0 | 27.5 | 32.8 |
| | | 02/15-09/16 | (24.4-24.6) | (29.2-29.5) | (34.8-35.1) | (22.8-23.3) | (27.2-27.8) | (32.4-33.2) |
| | | Round 18 | 25.6 | 30.4 | 36.0 | 22.5 | 27.1 | 32.0 |
| | | 10/16-05/18 | (25.5-25.8) | (30.2-30.5) | (35.8-36.1) | (22.1-22.9) | (26.8-27.4) | (31.5-32.4) |
| | | Round 19 | 27.1 | 32.2 | 37.7 | 22.2 | 28.3 | 34.5 |
| | | 06/18-05/19 | (26.9-27.3) | (32.0-32.3) | (37.6-37.9) | (21.7-22.8) | (27.7-29.0) | (33.9-35.0) |
| | Male | Round 16 | 27.6 | 32.3 | 37.8 | 26.3 | 30.4 | 36.2 |
| | | 07/13-01/15 | (27.5-27.8) | (32.1-32.5) | (37.5-38.0) | (26.1-26.5) | (30.2-30.7) | (35.8-36.6) |
| | | Round 17 | 28.8 | 33.0 | 37.9 | 27.5 | 31.8 | 36.9 |
| | | 02/15-09/16 | (28.6-29.0) | (32.8-33.2) | (37.7-38.1) | (27.3-27.8) | (31.5-32.1) | (36.5-37.2) |
| | | Round 18 | 29.5 | 34.1 | 39.4 | 27.1 | 31.8 | 36.8 |
| | | 10/16-05/18 | (29.3-29.6) | (33.9-34.2) | (39.2-39.5) | (26.7-27.5) | (31.5-32.2) | (36.5-37.2) |
| | | Round 19 | 30.6 | 35.4 | 40.7 | 28.3 | 31.8 | 36.0 |
| | | 06/18-05/19 | (30.4-30.9) | (35.2-35.6) | (40.5-40.9) | (27.9-28.7) | (31.4-32.3) | (35.4-36.7) |
| Inland | Female | Round 16 | 25.7 | 31.5 | 37.7 | 23.4 | 28.4 | 34.8 |
| | | 07/13-01/15 | (25.6-25.9) | (31.4-31.7) | (37.5-37.9) | (23.2-23.7) | (28.2-28.7) | (34.4-35.3) |
| | | Round 17 | 25.5 | 31.8 | 38.2 | 22.5 | 27.3 | 32.9 |
| | | 02/15-09/16 | (25.4-25.7) | (31.6-31.9) | (38.0-38.4) | (22.2-22.8) | (27.0-27.6) | (32.5-33.2) |
| | | Round 18 | 26.8 | 33.1 | 39.3 | 22.3 | 27.1 | 32.4 |
| | | 10/16-05/18 | (26.6-26.9) | (32.9-33.2) | (39.2-39.5) | (22.1-22.6) | (26.7-27.5) | (32.0-32.8) |
| | | Round 19 | 27.7 | 33.8 | 40.4 | 23.5 | 29.0 | 34.9 |
| | | 06/18-05/19 | (27.5-27.9) | (33.6-34.0) | (40.2-40.6) | (23.0-24.0) | (28.5-29.5) | (34.4-35.5) |
| | Male | Round 16 | 28.9 | 34.4 | 39.9 | 27.1 | 31.9 | 37.7 |
| | | 07/13-01/15 | (28.7-29.2) | (34.1-34.6) | (39.6-40.2) | (26.8-27.4) | (31.5-32.3) | (37.3-38.2) |
| | | Round 17 | 29.9 | 35.5 | 40.9 | 27.6 | 32.6 | 39.1 |
| | | 02/15-09/16 | (29.6-30.1) | (35.2-35.7) | (40.6-41.1) | (27.2-27.9) | (32.1-33.1) | (38.5-39.6) |
| | | Round 18 | 30.4 | 35.6 | 40.7 | 26.5 | 32.0 | 37.2 |
| | | 10/16-05/18 | (30.1-30.6) | (35.3-35.8) | (40.5-41.0) | (25.9-27.0) | (31.6-32.5) | (36.7-37.8) |
| | | Round 19 | 30.5 | 36.7 | 42.1 | 26.2 | 31.4 | 37.7 |
| | | 06/18-05/19 | (30.1-30.8) | (36.4-37.0) | (41.8-42.3) | (25.6-26.7) | (30.8-32.0) | (36.9-38.5) |

Supplementary Table S3: **Age profile of individuals with HIV and individuals with unsuppressed virus in the population by community type, gender and survey round, 2013-2019.** Age profiles are reported as posterior median estimates with 95% credible intervals in parentheses.

| Model | Round | PDI | EBFMI | ESS |
|---|---|---|---|---|
| HIV prevalence | 16 | 0.00 % | 0.96 | 6360 |
| in first-time participants | 17 | 0.00 % | 0.98 | 7598 |
| and all participants | 18 | 0.00 % | 1.00 | 7442 |
| | 19 | 0.00 % | 0.99 | 12589 |
| Viral suppression rates | 16 | 0.00 % | 0.99 | 3531 |
| among people with HIV | 17 | 0.00 % | 0.98 | 2234 |
| | 18 | 0.00 % | 0.99 | 2172 |
| | 19 | 0.00 % | 1.01 | 1647 |
| Prevalence of unsuppressed viraemia | 16 | 0.01 % | 0.96 | 3032 |
| in first-time participants | 17 | 0.00 % | 0.97 | 3277 |
| and all participants | 18 | 0.06 % | 0.98 | 5128 |
| | 19 | 0.04 % | 0.96 | 3044 |

Supplementary Table S4: **Convergence diagnostics for all models**. PDI=Proportion of Divergent Iterations, EBFMI= Effective Bayesian Fraction of Missing Information, ESS= Minimum effective sample size for the model parameters.

#### Supplementary Figures

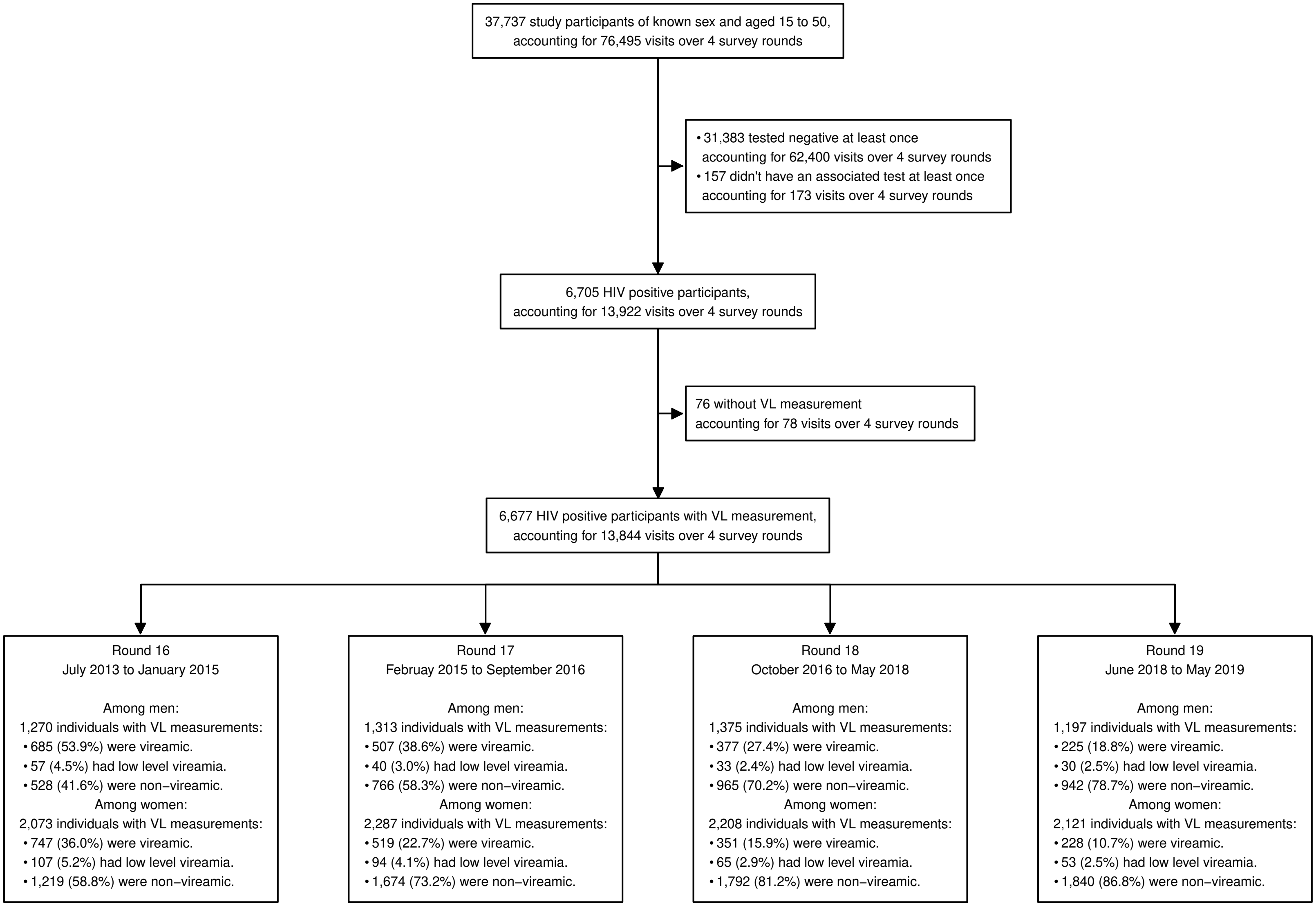


Supplementary Figure S1: **Data flowchart.**

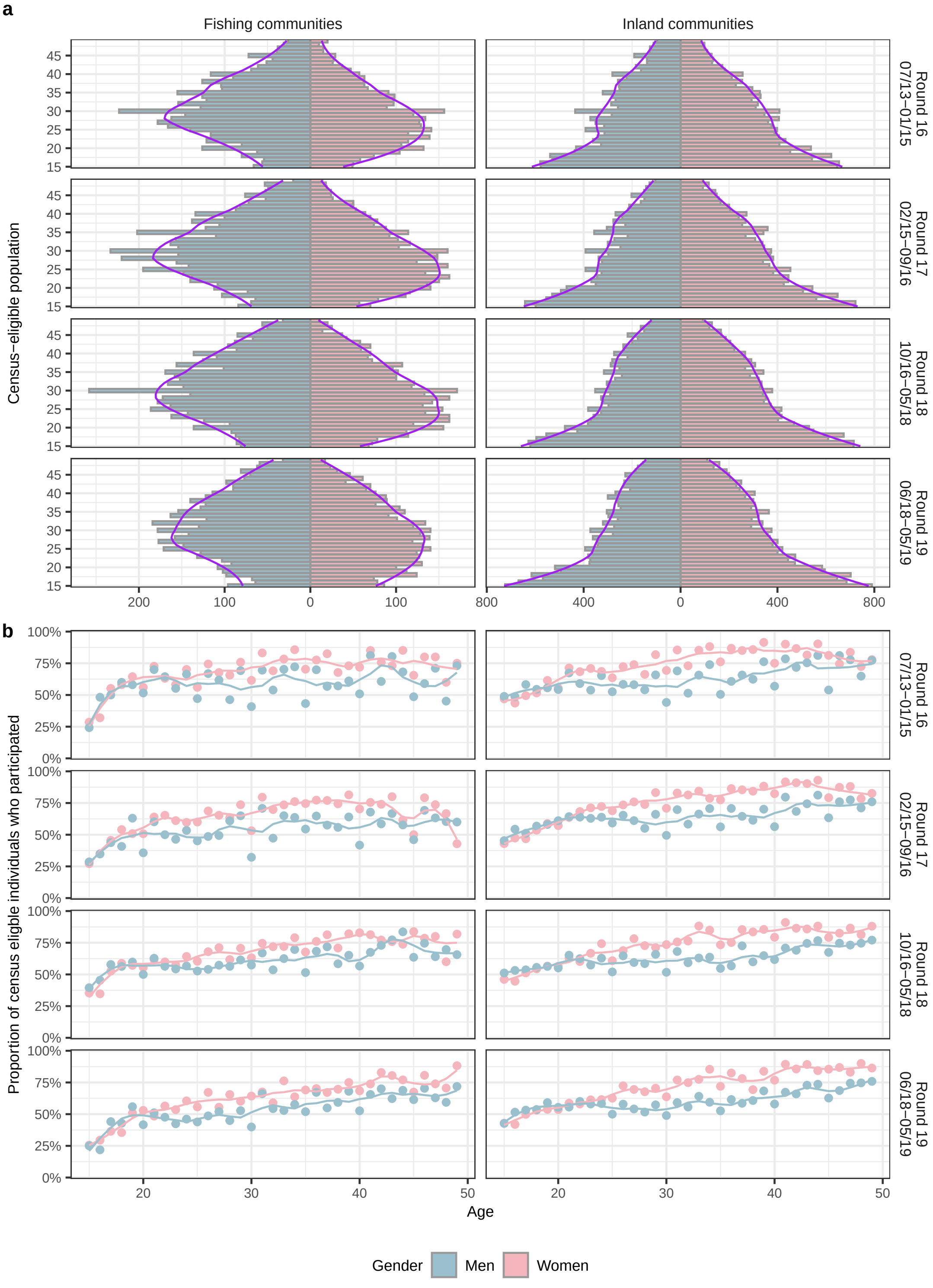


Supplementary Figure S2: **RCCS population size estimates and participation rates, 2013-2019 (a)** Age- and gender-specific population sizes reported by households heads during the census preceeding each survey round (bars), overlayed with smooth maximum likelihood population size estimates obtained with LOESS (purple line). **(b)** Age- and gender-specific empirical survey participation rates in the census-eligible populations (dots), overlayed with smooth maximum likelihood estimates obtained with LOESS (lines).

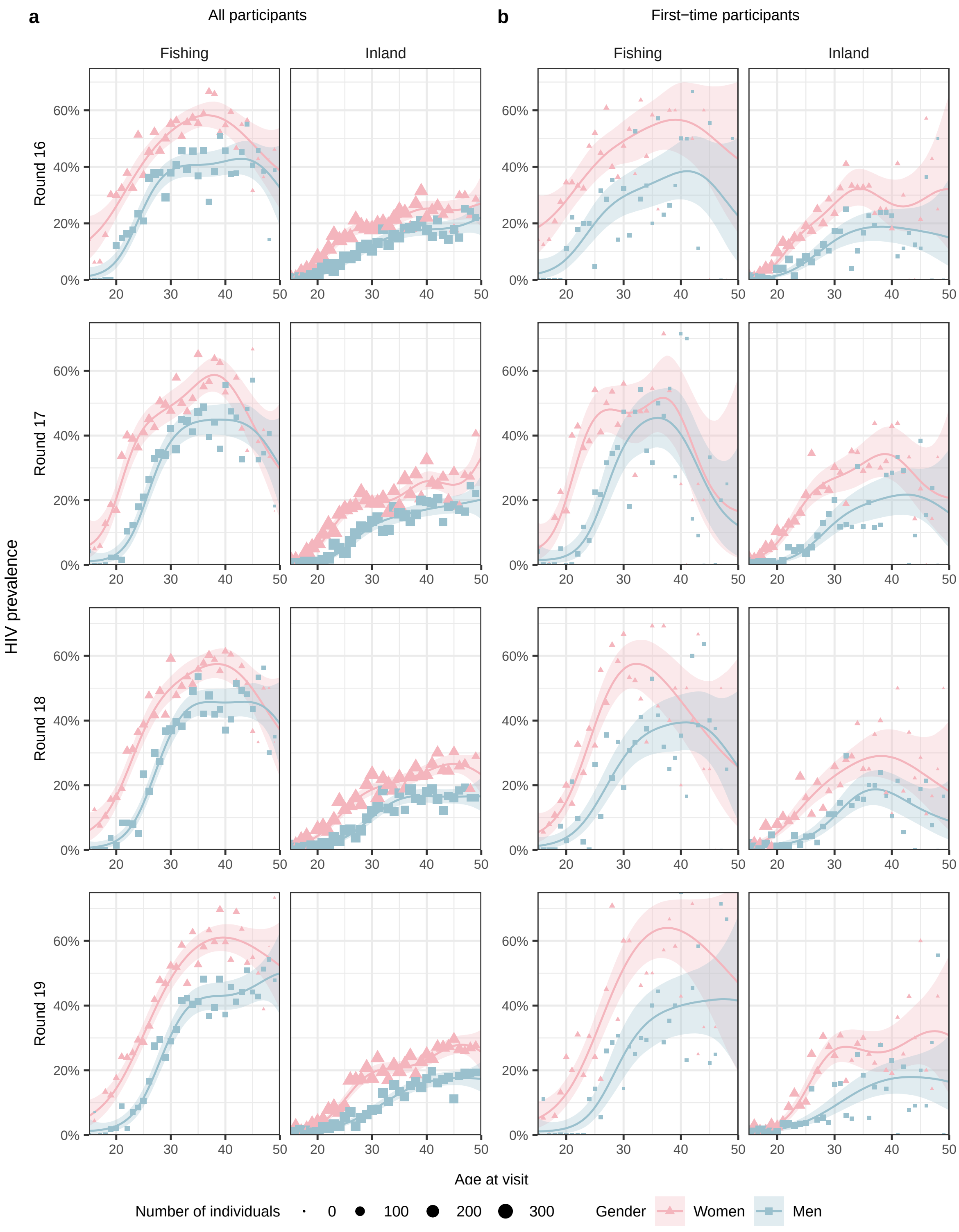


Supplementary Figure S3: **Age- and gender-specific HIV prevalence estimates among study participants and first-time participants, 2013-2019. (a)** Posterior median estimates of the proportion of all participants in each age and gender group who have HIV (lines) and 95% CrI (ribbons), shown along empirical proportions (squares and triangles for men and women respectively). Participant numbers in each stratum are indicated by the size of squares or triangles. **(b)** Same for the proportion of first-time participants in each age and gender group who have HIV.

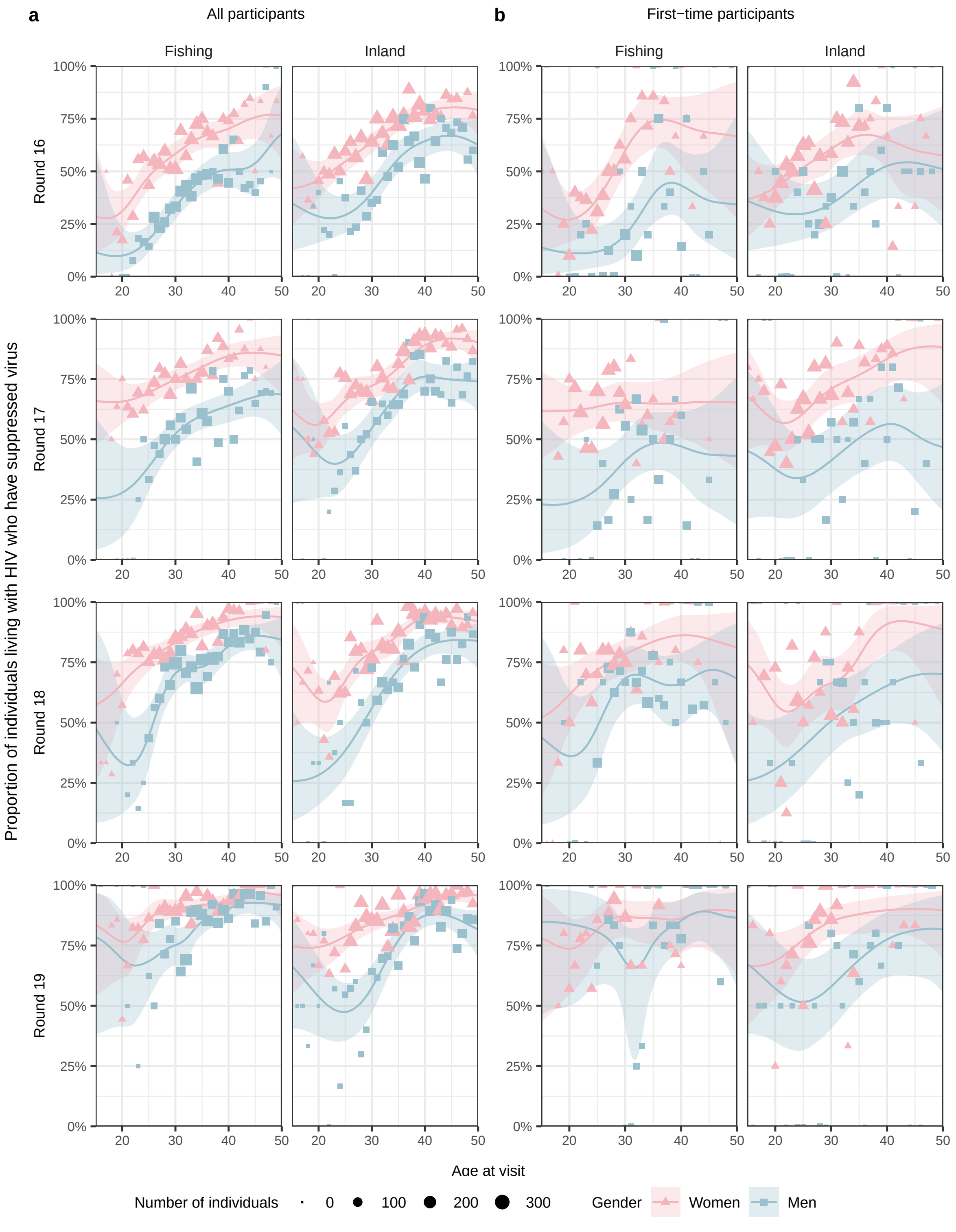


Supplementary Figure S4: **Age- and gender-specific viral load suppression rate estimates among study participants and first-time participants with HIV, 2013-2019. (a)** Posterior median estimates of the proportion of all participants with HIV in each age and gender group who have suppressed virus (lines) and 95% CrI (ribbons), shown along empirical proportions (squares and triangles for men and women respectively). Participant numbers in each stratum are indicated by the size of squares or triangles. **(b)** Same for the proportion of first-time participants with HIV in each age and gender group who have suppressed virus.

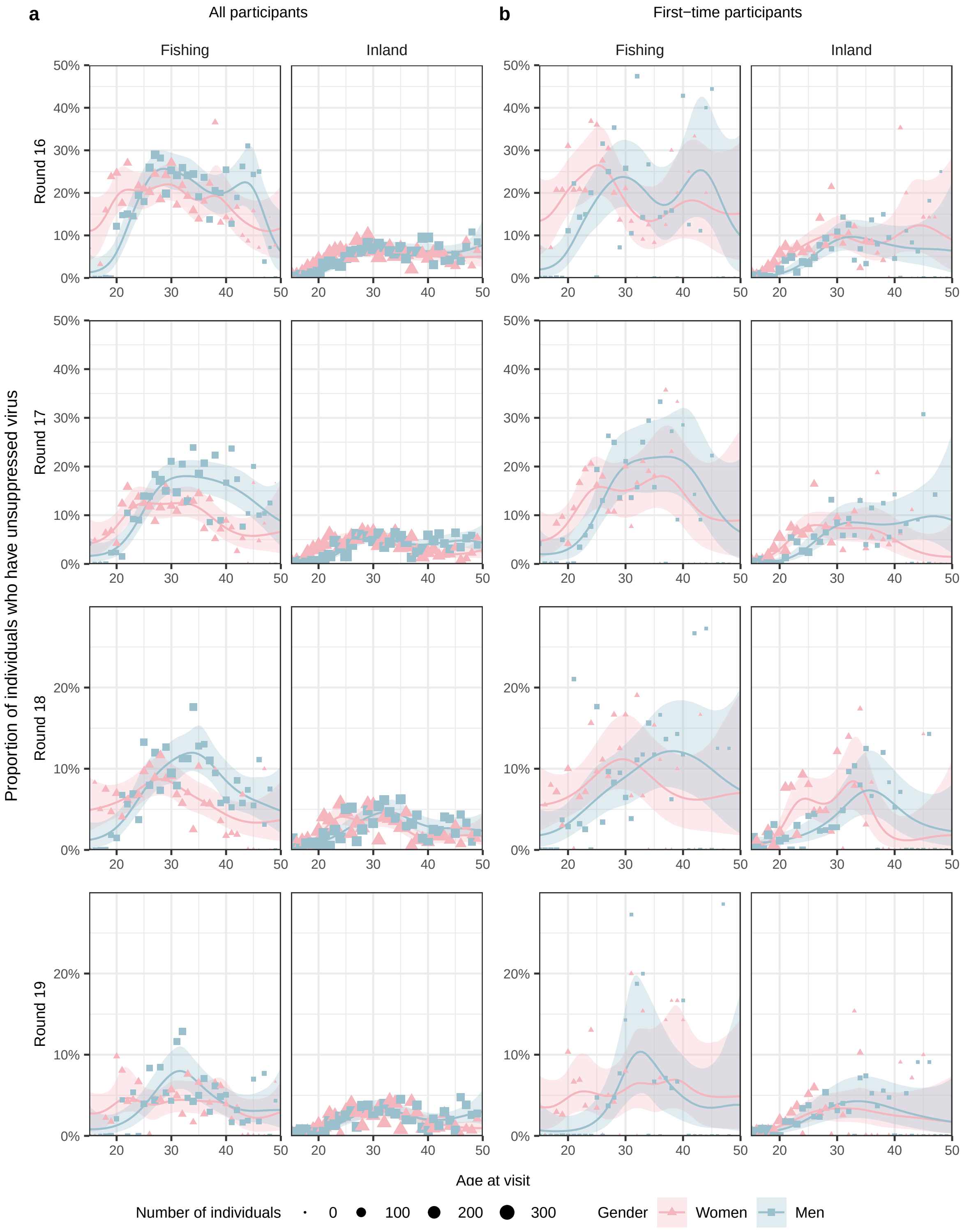


Supplementary Figure S5: **Age- and gender-specific viral load suppression rate estimates of among census-eligible people with HIV.** Posterior median estimates of the proportion of census-eligible people with HIV in each age and gender group exhibiting viraemia (lines) and 95% CrI (ribbons), shown along empirical proportions (squares and triangles for men and women respectively). Participant numbers in each stratum are indicated by the size of squares or triangles.

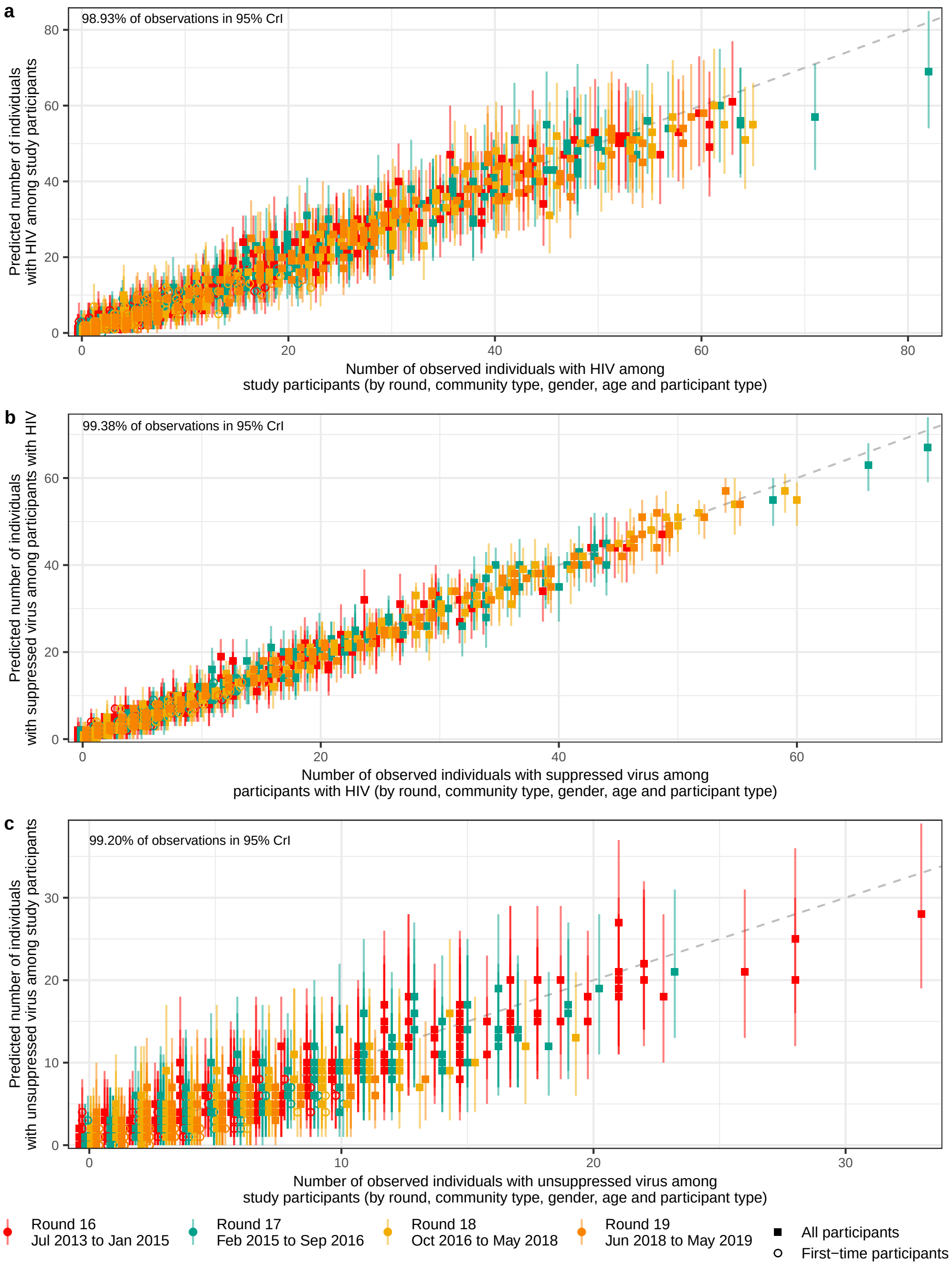


Supplementary Figure S6: **Consistency of fitted models with available data.** **(a)** Posterior predictive median estimates of the number of individuals with HIV among participants (dot) are plotted on the y-axis versus the observed number in each survey round-community type-gender-age-participant type stratum, along with 95% posterior predictive intervals (errorbars). **(b)** Posterior predictive median estimates of the number of individuals suppressed virus among participants with HIV (dot) are plotted on the y-axis versus the observed number in each survey round-community type-gender-age-participant type stratum, along with 95% posterior predictive intervals (errorbars). **(c)** Posterior predictive median estimates of the number of individuals suppressed virus among participants (dot) are plotted on the y-axis versus the observed number in each survey round-community type-gender-age-participant type stratum, along with 95% posterior predictive intervals (errorbars).

### S1 Extended Methods

#### S1.1 Population size and participation rates

To account for population structures, we used RCCS's census data which reported census-eligible population sizes by community type, survey rounds, gender and 1-year age bands. The ages reported by household heads during the census reflected grouping patterns towards ages which were multiples of 5. As such, we obtained smoothed population sizes $\hat{N}_{g,a}$ for gender $g$ across ages $a \in \mathcal{A} \in \{15, 16, \ldots, 49\}$ years, stratifying by round and community type using locally weighted running line smoothers (LOESS) as implemented in the `R` package `stats` version `4.2.2` with `span=0.5`. We analogously obtained smooth estimates of participation rates $\hat{\theta}^{\text{loess}}_{g_a}$ of census-eligible populations in each community type, survey round, gender and 1-year age band stratum. Here, the LOESS span argument was set to `span=0.25`. The data and resulting fits are illustrated in Supplementary Figure S2.

#### S1.2 Population-level HIV prevalence estimates by age and gender

Our primary objective was to obtain smooth estimates of the proportion of the census-eligible population in each community type, survey round, gender and 1-year age band stratum who have HIV, and similarly to obtain smooth estimates of the age and gender profile of the census-eligible population with HIV in each community type and survey round. We obtained smooth age-specific estimates independently for each gender, survey round and community type combination, and below drop notation relevant to gender, survey round and community type. Throughout, we fitted models to population-based HIV positive test data from both all participants (denoted by $(r = 1)$), and first-time participants (denoted by $(r = 2)$). We nonetheless treated all data points independent and did not consider first-time and repeat participants because in young age groups all participants were first-timers and this posed challenges in obtaining robust non-parametric prevalence estimates under this alternative approach. For each participant type $r$ and age group $a$, we denote the number of study participants with $Z_{a,r}$ and the subset of participants with HIV by $Y_{a,r}$. We modelled age-specific HIV prevalence rates with the non-parametric Bayesian logit model [4].

$$
\begin{aligned}
Y_{a,r} &\sim \text{Binomial}\left(Z_{a,r}, \pi^{\text{hiv}}_{a,r}\right) && \text{(1a)}\\
\text{logit}(\pi^{\text{hiv}}_{a,r}) &= \nu_r + f_r(a) && \text{(1b)}\\
\nu_r &\sim \text{Normal}\left(0, 10^2\right) && \text{(1c)}\\
f_r &\sim \text{Gaussian-Process}\left(0, k_{\sigma,\rho}\right) && \text{(1d)}\\
\sigma &\sim \text{Half-Normal}\left(0, 10^2\right) && \text{(1e)}\\
\rho &\sim \text{Inverse-Gamma}\left(\rho_{\text{shape}}, \rho_{\text{scale}}\right), && \text{(1f)}
\end{aligned}
$$

where $a \in \mathcal{A}$, and $r \in \{1, 2\}$. Age-specific HIV prevalence in all participants and first-time participants was modelled on the logit scale with fixed, independent baseline effects $\nu_r$ plus 1-dimensional Gaussian processes priors $f_r(a)$ specified by squared exponential kernels $k_{\sigma,\rho}$ with variance parameter $\sigma$ and length-scale $\rho$. Age inputs were translated to the interval $[1, 35]$. The hyperparameters $\rho_{\text{shape}}$ and $\rho_{\text{scale}}$ where chosen to address identifiability issues relating to minimum and maximum covariate distances, and set such that 99% prior mass was contained within $[1, 35]$, i.e. $\rho_{\text{shape}} = 2.21$ and $\rho_{\text{scale}} = 7.05$ [1].

Model (1) was fitted repeatedly to all gender, survey round and community type combinations with the `R` package `cmdstanr` version `0.5.3`, using four separate HMC chains with 500 warmup iterations and 10,000 sampling iterations each [3, 2]. Convergence and mixing diagnostics are reported in Supplementary Table S4, and the model fitted all the available data well (Supplementary Figs. S3,S6).

From the fitted models, we generated smooth age-specific estimates of the proportion of the census-eligible population in each community type (denoted by $c$), survey round (denoted by $t$), gender $g$ and 1-year age band $a$ stratum who have HIV with

$$\pi^{\text{hiv-pop}}_{c,t,g,a} = \hat{\theta}^{\text{loess}}_{c,t,g,a} \times \pi^{\text{hiv}}_{c,t,g,a,r=1} + (1 - \hat{\theta}^{\text{loess}}_{c,t,g,a}) \times \pi^{\text{hiv}}_{c,t,g,a,r=2}, \tag{2}$$

where $\hat{\theta}^{\text{loess}}_{c,t,g,a}$ are the smooth participation rate estimates and $\pi^{\text{hiv}}_{c,t,g,a,r=1}$ are the posterior HIV prevalence estimates among all participants and $\pi^{\text{hiv}}_{c,t,g,a,r=2}$ are the posterior HIV prevalence estimates among first-time participants from model (1). In Eq. (2), we assume HIV prevalence in the subpopulations who did not participate is the same as HIV prevalence among first-time participants.

#### S1.3 Age and gender profiles of people living with HIV

To investigate the age and gender profile of people with HIV in each community type (denoted by $c$) and survey round (denoted by $t$), we first estimated the number of people with HIV of gender $g$ and age $a$ as:

$$I_{c,t,g,a} = \hat{N}_{c,t,g,a} \times \pi^{\text{hiv-pop}}_{c,t,g,a}, \tag{3}$$

where $\pi^{\text{hiv-pop}}_{c,t,g,a}$ are from Eq. (2), $\hat{N}_{c,t,g,a}$ are the LOESS-smoothed population size estimates. Next, we generated the age and gender profile of people with HIV in each community type $c$ and survey round $t$, by normalising the age and gender specific number of individuals HIV by the total over all age groups and genders:

$$\gamma^{\text{hiv}}_{c,t}(a,g) = I_{c,t,g,a} \Big/ \Big( \sum_{a' \in \mathcal{A}, g' \in \mathcal{G}} I_{c,t,g',a'} \Big), \tag{4}$$

where $\mathcal{G} = \{\text{men}, \text{women}\}$. In Eq. (4), the profile sums to 1 over all age and gender groups in each subpopulation specified by community type and survey round. The age and gender profile of HIV prevalence also accounts for population structure between men and women, as for example in fishing communities the gender ratio is far from 50:50.

#### S1.4 Population-level prevalence of viraemia estimates by age and gender among PLHIV

Our objective was to obtain smooth estimates of the proportion of the census-eligible population with HIV in each community type, survey round, gender and 1-year age band stratum who had suppressed virus. We obtained smooth age-specific estimates independently for each gender, survey round and community type combination using the same model as for estimating population-level HIV prevalence. Specifically, and suppressing again notation for community type, survey round and gender, we fitted model (1) to the number $\tilde{Z}_{a,r}$ of study participants with HIV in age band $a$ and of participant type $r$, and the subset $\tilde{Y}_{a,r}$ of participants with HIV in age band $a$ and of participant type $r$ who had suppressed virus. Convergence and mixing diagnostics are reported in Supplementary Table S4, and the model fitted all the available data well

(Supplementary Figs. S4,S6).

From the fitted model, we obtained smooth age-specific estimates $\pi^{\text{sup-pop}}_{c,t,g,a}$ of the proportion of the census-eligible population with HIV in each community type (denoted by $c$), survey round (denoted by $t$), gender $g$ and 1-year age band $a$ stratum who had suppressed virus in analogy to Eq. (2).

#### S1.5 Age and gender profile of people exhibiting viraemia

Finally, we sought to obtain smooth estimates of the proportion of the census-eligible population in each community type, survey round, gender and 1-year age band stratum exhibiting viraemia, and similarly to obtain smooth estimates of the age and gender profile of the census-eligible population exhibiting viraemia in each community type and survey round. We again obtained smooth age-specific estimates independently for each gender, survey round and community type combination using the same model as for estimating population-level HIV prevalence. Specifically, and suppressing again notation for community type, survey round and gender, we fitted model (1) to the number $Z_{a,r}$ of study participants in age band $a$ and of participant type $r$, and the subset $\tilde{Z}_{a,r} - \tilde{Y}_{a,r}$ of participants exhibiting viraemia in age band $a$ and of participant type $r$. Convergence and mixing diagnostics are reported in Supplementary Table S4, and the model fitted all the available data well (Supplementary Figs. S5,S6).

From the fitted model, we obtained smooth age-specific estimates $\pi^{\text{vir-pop}}_{c,t,g,a}$ of the proportion of the census-eligible population in each community type (denoted by $c$), survey round (denoted by $t$), gender $g$ and 1-year age band $a$ stratum who had suppressed virus in analogy to Eq. (2).

Similarly, we obtained estimates of the number of individuals exhibiting viraemia $V_{c,t,g,a}$ analogously to Eq. (3) and computed the age profile $\gamma_{c,t}(a)$ of individuals exhibiting viraemia in each community type and survey round as in Eq. (4), replacing $I_{c,t,g,a}$ by $V_{c,t,g,a}$. Namely, the number $V_{c,t,g,a}$ of individuals exhibiting viraemia in community type $c$, at round $t$ of gender $g$ and age $a$ was defined as:

$$V_{c,t,g,a} = \hat{N}_{c,t,g,a} \times \pi^{\text{vir-pop}}_{c,t,g,a}, \tag{5}$$

while the age and gender profile of individuals exhibiting viraemia in each community type $c$ and survey round $t$ was defined as:

$$\gamma^{\text{vir}}_{c,t}(a, g) = V_{c,t,g,a} \Big/ \Big( \sum_{a' \in \mathcal{A}, g' \in \mathcal{G}} V_{c,t,g',a'} \Big). \tag{6}$$

#### S1.6 Age quantiles for burden profiles by gender

We decided to capture and summarize changes in the burden profiles of PLHIV by calculating the $p$ quantiles $q^{\text{hiv}-p}_{t,c,g}$ of the age profiles by survey round $t$, community type $c$, and gender $g$ for $p \in \{25\%, 50\%, 75\%\}$. Specifically, for every Monte Carlo iteration, we used the profiles defined in Equation 4 to define a continuous probability distribution $g^{\text{hiv}}_{c,t,g}$ over the ages for every gender, and evaluated the quantiles for these distributions. We simply assumed $g^{\text{hiv}}_{c,t,g}$ to be a step function such that for every age integer $a$, the distribution is constant on $[a, a + 1)$, and equals $\gamma^{\text{hiv}}_{t,c}(g, a)$. Then, for every $p \in \{25\%, 50\%, 75\%\}$, we solve:

$$p = \int_{15}^{q_{t,c,g}^{\mathrm{hiv}-p}} g_{c,t,g}^{\mathrm{hiv}}(x)dx, \tag{7}$$

in terms of $q_{t,c,g}^{\mathrm{hiv}-p}$.

We analogously calculated $p$ quantiles $q_{t,c,g}^{\mathrm{vir}-p}$ for the age profile of people exhibiting viraemia. Specifically, we defined a continuous probability distribution $g_{c,t,g}^{\mathrm{vir}}$ from the burden profile defined in Equation 6, and solved an analogous equation to Equation 7 for every community type $c$, survey round $t$, and gender $g$.

### References

[1] Michael Betancourt. *Robust Gaussian Process Modeling.* URL: https://betanalpha.github.io/assets/case_studies/gaussian_processes.html (visited on 10/10/2023).

[2] Bob Carpenter et al. "Stan: A Probabilistic Programming Language". In: *Journal of Statistical Software* 76 (Jan. 2017), pp. 1–32. ISSN: 1548-7660. DOI: 10.18637/jss.v076.i01. URL: https://doi.org/10.18637/jss.v076.i01 (visited on 10/20/2023).

[3] Jonah Gabry, Rok Češnovar, and Andrew Johnson. *Cmdstanr: R Interface to 'CmdStan'*. Manual. 2023.

[4] Mélodie Monod et al. "Longitudinal Population-Level HIV Epidemiologic and Genomic Surveillance Highlights Growing Gender Disparity of HIV Transmission in Uganda". In: *Nature Microbiology* 9.1 (Dec. 2023), pp. 35–54. ISSN: 2058-5276. DOI: 10.1038/s41564-023-01530-8. URL: https://www.nature.com/articles/s41564-023-01530-8 (visited on 02/20/2024).
